# The global landscape of susceptibility to orthopoxviruses: The legacy of smallpox vaccination

**DOI:** 10.1101/2022.07.29.22278217

**Authors:** Juliana C. Taube, Eva C. Rest, James O. Lloyd-Smith, Shweta Bansal

## Abstract

**Background:** More than four decades after the eradication of smallpox, the ongoing 2022 monkeypox outbreak and increasing transmission events of other orthopoxviruses necessitate a deeper understanding of the global distribution of susceptibility to orthopoxviruses, as shaped by the landscape of smallpox vaccination pre-eradication.

**Methods:** We characterize the fine-scale global spatial landscape of orthopoxvirus vulnerability based on geographical heterogeneity in demography and past smallpox vaccination program features, including vaccination coverage and cessation dates. For the United States, we also consider the role of immigration in shaping the landscape of protection.

**Findings:** We find significant global spatial heterogeneity in the landscape of orthopoxvirus susceptibility, with monkeypox susceptibility ranging from near 57% to near 96% within admin-1 regions globally, with negligible uncertainty in most regions. We identify that this variation is driven largely by differences in sub-national demography.

**Interpretation:** The legacy of smallpox eradication can be observed in the current landscape of susceptibility for orthopoxviruses, including monkeypox. The strength and longevity of the smallpox vaccination campaign in each nation shape the epidemiological landscape today and reveal significant geographic variation in vulnerability. Our work alerts public health decision-makers to non-endemic regions that may be at greatest risk in the case of widespread and sustained transmission in the 2022 monkeypox outbreak and highlights the importance of demography and fine-scale spatial dynamics in predicting future public health risks from orthopoxviruses.

**Funding:** Research reported in this publication was supported by the National Institutes of Health under award number R01GM123007 (SB) and National Science Foundation DEB-1557022 (JOL-S).

**Research in Context:** 

**Evidence before this study:** We searched the Red Book, WHO eradication documents, *Bulletin of the World Health Organization* and *Morbidity and Mortality Weekly Report* digital archives, and published literature and reports via Google Scholar and PubMed for data on smallpox vaccination coverage rates and cessation dates for each country. Search terms included: “smallpox vaccination cessation”,”end of smallpox vaccination”, “stop smallpox vaccination”, “smallpox vaccination coverage”, “smallpox scar surveys”, and “smallpox serum surveys”, combined with each country name. While the Red Book covers much of the smallpox eradication efforts in Africa, Asia, and South America, information on countries in Europe, Central America, the Middle East, and Oceania was sparse and scattered.

**Added value of this study:** We characterize the fine-scale global spatial landscape of orthopoxvirus vulnerability based on geographical heterogeneity in demography and past smallpox vaccination program features, including vaccination coverage and cessation dates. We find significant spatial heterogeneity in orthopoxvirus susceptibility, driven in large part by age structure, specifically what proportion of the population in a region was born before smallpox vaccination cessation. We contribute an open (and living) database of all subnational susceptibility estimates and uncertainties as an immediate resource for the global health community working on the monkeypox outbreak.

**Implications of all of the available evidence:** Our findings highlight the need to consider spatial clustering of susceptible individuals and the importance of fine-scale spatial analysis in light of increased risk of orthopoxvirus emergence. In the event that transmission becomes widespread during the 2022 global monkeypox outbreak, our vulnerability map can inform public health efforts on identifying non-endemic regions and age cohorts at greatest risk, allocation of scarce vaccine supplies, and predicting transmission dynamics in concert with data on contact patterns, mobility and real-time prevalence.

## Introduction

More than four decades after the landmark eradication of smallpox, the ongoing 2022 outbreak of monkeypox in over 70 non-endemic countries, threatens global health. The spatial distribution of sustained and widespread monkeypox transmission, if the outbreak is not contained, will be shaped by heterogeneity in the smallpox immune landscape, due to cross-protection conferred by the *Vaccinia*-based smallpox vaccine. Multiple factors influence this immune landscape, including country-specific smallpox endemicity and elimination timelines, protocols for routine smallpox vaccination, and demographic and migration patterns post-eradication. However, global estimates of the current population immunized against smallpox are unavailable, limiting our ability to predict large-scale transmission dynamics of the current monkeypox outbreak or of potential future outbreaks of other *Orthopoxvirus* genus viruses. We seek to characterize the current global spatial heterogeneity in orthopoxvirus susceptibility as driven by geographic variation in demography, cessation dates for routine smallpox vaccination, and vaccination coverage.

Smallpox afflicted humans for millennia and remains to date the only successfully eradicated human disease. The World Health Organization (WHO) committed to an intensified global smallpox eradication campaign in 1967, particularly focusing on countries with high rates of endemic smallpox across Africa, Asia, and South America.1 Although the WHO initially set a target vaccination rate of 80% worldwide, novel vaccination strategies helped eliminate smallpox country-by-country while also introducing spatial heterogeneity in vaccination coverage based on the vaccination campaign of each nation. The jet injector enabled mass vaccination efforts, the bifurcated needle stretched vaccine stocks further, and the use of targeted ring vaccination allowed for successful elimination at lower vaccination rates.^1^ Progress towards elimination in each country was tracked via scar surveys for vaccination and pockmark surveys for infection (both considered validated measures of immunity).^1, 2^ In 1977, just ten years into the intensified eradication campaign, the last naturally-occurring case of smallpox occurred in Somalia; the WHO declared smallpox eradicated in 1980.^3^ There have been no naturally-occurring smallpox cases since, but the use of smallpox as a biological weapon remains a risk.

Monkeypox is an orthopoxvirus closely related to but less severe than smallpox. Unlike smallpox, monkeypox was historically geographically limited to central and western Africa with relatively small outbreaks of zoonotic origin.^4^ Studies during the intensified WHO monkeypox surveillance period in the 1980s found that previous smallpox vaccination led to reduced secondary attack rates and reduced symptom severity upon monkeypox infection, estimating 85% effectiveness of the *Vaccinia*-based smallpox vaccine against monkeypox infection and disease.^5–7^ More recent studies support these findings, reporting continued evidence that smallpox vaccination prevented clinical monkeypox infection with >80% effectiveness and reduced infection severity in cases that did arise.^8–10^

Monkeypox outbreaks have risen in frequency and size since routine smallpox vaccination ended and reports of zoonotic transmission of cowpox and buffalopox, two other members of the *Orthopoxvirus* genus which infect humans, have also increased.^8, 11–13^ Since the *Vaccinia*-based smallpox vaccine confers some level of immunity against almost all human orthopoxviruses, we expect these other viruses to benefit from the growth in unvaccinated populations.^14, 15^ Mapping global heterogeneity in smallpox vaccination coverage and cessation is thus important to accurately understand the populations most vulnerable to sustained transmission arising from the 2022 monkeypox outbreak, a smallpox biological attack, or future emerging orthopoxvirus outbreaks. However, considering these processes at large spatial scales obscures important geographical heterogeneity that can drive disease transmission dynamics, including potentially high-risk areas where targeted interventions may be needed, and necessitates a fine-scale approach to characterizing the orthopoxvirus susceptibility landscape.^16–18^

Here, we characterize the current global spatial landscape of orthopoxvirus susceptibility based on geographical heterogeneity in demography and past smallpox vaccination efforts. We consider age distributions of populations, smallpox vaccination coverage (by age, where available) and smallpox vaccination cessation dates at the national and sub-national (admin-1 or finer) levels. For the U.S., we also consider the role of immigration in shaping the landscape of protection. We characterize uncertainties in these estimates, and analyze the role of each demographic and vaccination campaign characteristic. While other factors, like contact patterns, remain critical to predicting monkeypox transmission, our work maps the current global landscape of susceptibility to monkeypox and other orthopoxviruses, providing key information to guide public health decision-making for the evolving 2022 outbreak and beyond.

## Results

We develop high-resolution global and U.S. maps of current population susceptibility against monkeypox based on sub-national and age-specific estimates of demography and vaccination efforts, and find substantial variation across regions with susceptibility levels ranging from near 50% to near 100% (Figures 1A, 2A). We perform an uncertainty analysis to understand the robustness of our estimates and find that 82% of admin-1 region estimates differ from the original values by less than 1%, and only 1% of regions differ by 5 to 9% (Figure S1). To investigate the possible effect of waning immunity, we also consider the sensitivity of our estimates to different levels of cross-protection from smallpox vaccine towards monkeypox and find that our estimates vary little with this choice (< 3 percentage points, Figure S2). To additionally characterize the susceptibility landscape for other orthopoxviruses, we consider values of smallpox vaccine effectiveness against *Variola major* and *Variola minor* (Figure S3). We do not include the effect of natural immunity to smallpox in our estimates because prevalence was low during the 1970s even in the last smallpox endemic countries (Figures S4, S5, S6). Lastly, we compare our projected estimates of population susceptibility to field data from recent smallpox scar surveys and antibody studies from five countries, and find that our model estimates are consistent with the empirical estimates (Figure S7).

**Figure 1:**
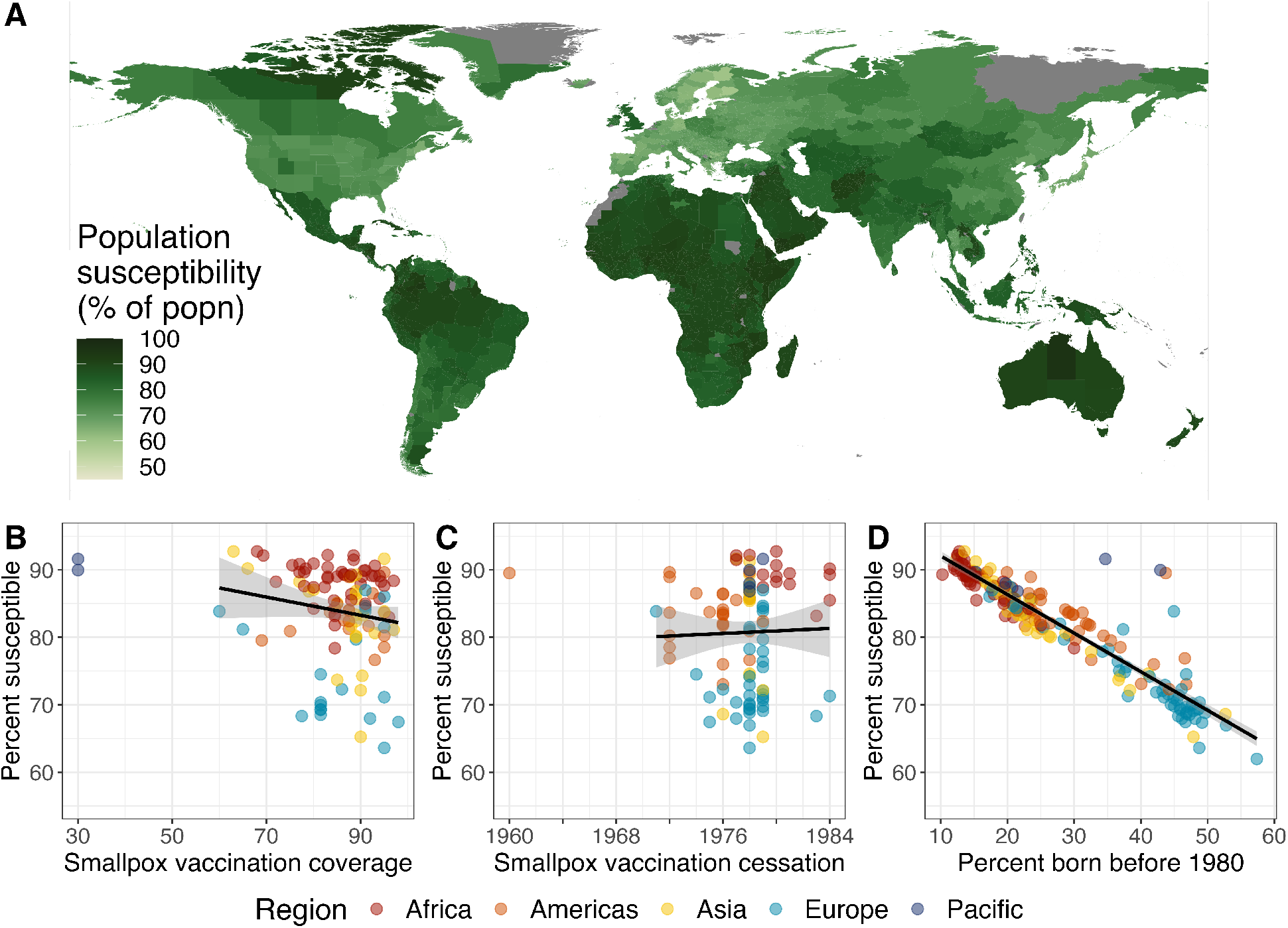
The global landscape of monkeypox susceptibility. (A) Population susceptibility to monkeypox infection at the admin-1 scale globally. (B-D) Relationship between the average national population susceptibility to monkeypox and (B) average national smallpox vaccination coverage near cessation, (C) the year that routine smallpox vaccination ceased within the country, and (D) the proportion of the country’s current population that was born before 1980. The black line is based on a linear regression of the data, not including outliers. Outlier countries are Australia and New Zealand (low coverage), Cuba (early cessation), and the United Kingdom (low coverage). Vaccination coverage and cessation date are not predictive of susceptibility on their own, while the proportion of the population born before 1980 is.

### Significant spatial heterogeneity exists in orthopoxvirus susceptibility

We find high spatial heterogeneity in the landscape of orthopoxvirus vulnerability, with admin-1-level population susceptibility ranging from 57 to 96% globally, and varying by up to 8% nationally (Figure 1A, S8). Parts of Finland, Bulgaria, Monaco, Japan, and Sweden have the lowest levels of susceptibility, while some of the most susceptible regions are in Australia, Yemen, Colombia, Guinea-Bissau, and Ethiopia. Significant sub-national heterogeneity can be observed in many areas but is particularly evident in large countries such as India, China, Brazil, and the U.S. (Figures 1A, S8). Given that central and western Africa are common locations for monkeypox spillover, their high levels of susceptibility are notable. A similar landscape can be observed for smallpox (*Variola major* and *Variola minor*) susceptibility (Figure S3).

In Figure 1B-D, we demonstrate the role of demographic and vaccination factors in shaping the susceptibility landscape. We find that a nation’s current demography (as measured by the proportion of the population born before smallpox eradication in 1980) is highly predictive of monkeypox vulnerability (Pearson’s correlation, *ρ* = −0·93). Additionally, while smallpox vaccination coverage near cessation is not predictive on its own, it is correlated to susceptibility in countries with older populations (e.g., in Europe and Asia).

Given finer-scale demographic and vaccination data available in the U.S., we estimate monkeypox population immunity at the PUMA level (census geographical areas with populations of at least 100,000, Figure 2A). There is high heterogeneity in monkeypox protection across the U.S. with PUMA-level susceptibility levels ranging from 46% to 90%. Regions in Florida and Arizona are the least susceptible, while a number of regions in Utah are the most susceptible. In Figure 2B-D, we find that similar to the global landscape, demography is highly predictive of current monkeypox susceptibility (*ρ* = − 0 · 95). Additionally, variation in smallpox vaccination coverage is predictive of susceptibility (*ρ* = 0 · 54), but the proportion of the population born outside the U.S. is not significantly associated.

**Figure 2:**
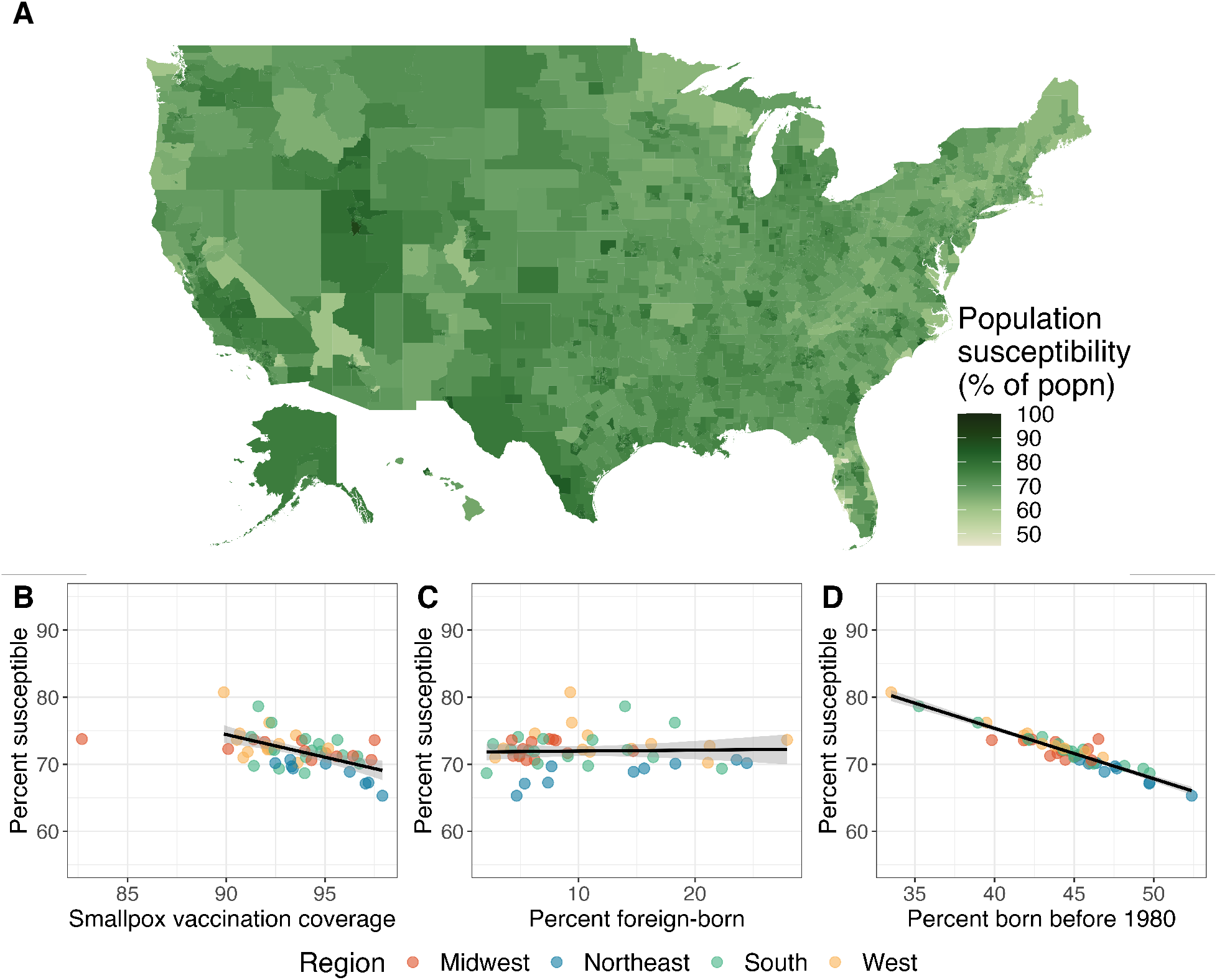
The national landscape of monkeypox susceptibility in the U.S. (A) Population susceptibility to monkeypox infection within the United States across geographic regions with at least 100,000 individuals. (B-D) Relationship between the average U.S. state population susceptibility to monkeypox and state-specific smallpox vaccination coverage at cessation, (C) the proportion of the state’s population that is foreign-born, and (D) the proportion of the state’s current population that was born before 1980. The black line is based on a linear regression of the data, not including outliers. The state of Michigan is the only outlier with an usually low vaccination coverage.

### Demography is a key driver of spatial heterogeneity in monkeypox vulnerability

To tease out the factors driving the observed spatial heterogeneity in the monkeypox immune landscape, we compare estimates under four counterfactual scenarios: (a) simultaneous cessation (i.e., all countries share a routine smallpox vaccination cessation date of 1984); (b) universal vaccination efforts (i.e., all countries achieve 100% vaccination coverage before cessation); (c) homogeneous demography (i.e., all countries and regions share the global average age distribution); and (d) no immigration (i.e., the U.S. has no foreign-born individuals and all remaining individuals are subject to U.S. smallpox vaccination efforts). In Figure 3, we summarize the impact of each scenario relative to the empirical estimates (summarized in Figures 1A and 2A). All four counterfactuals highlight significant differences in protection under different conditions.

**Figure 3:**
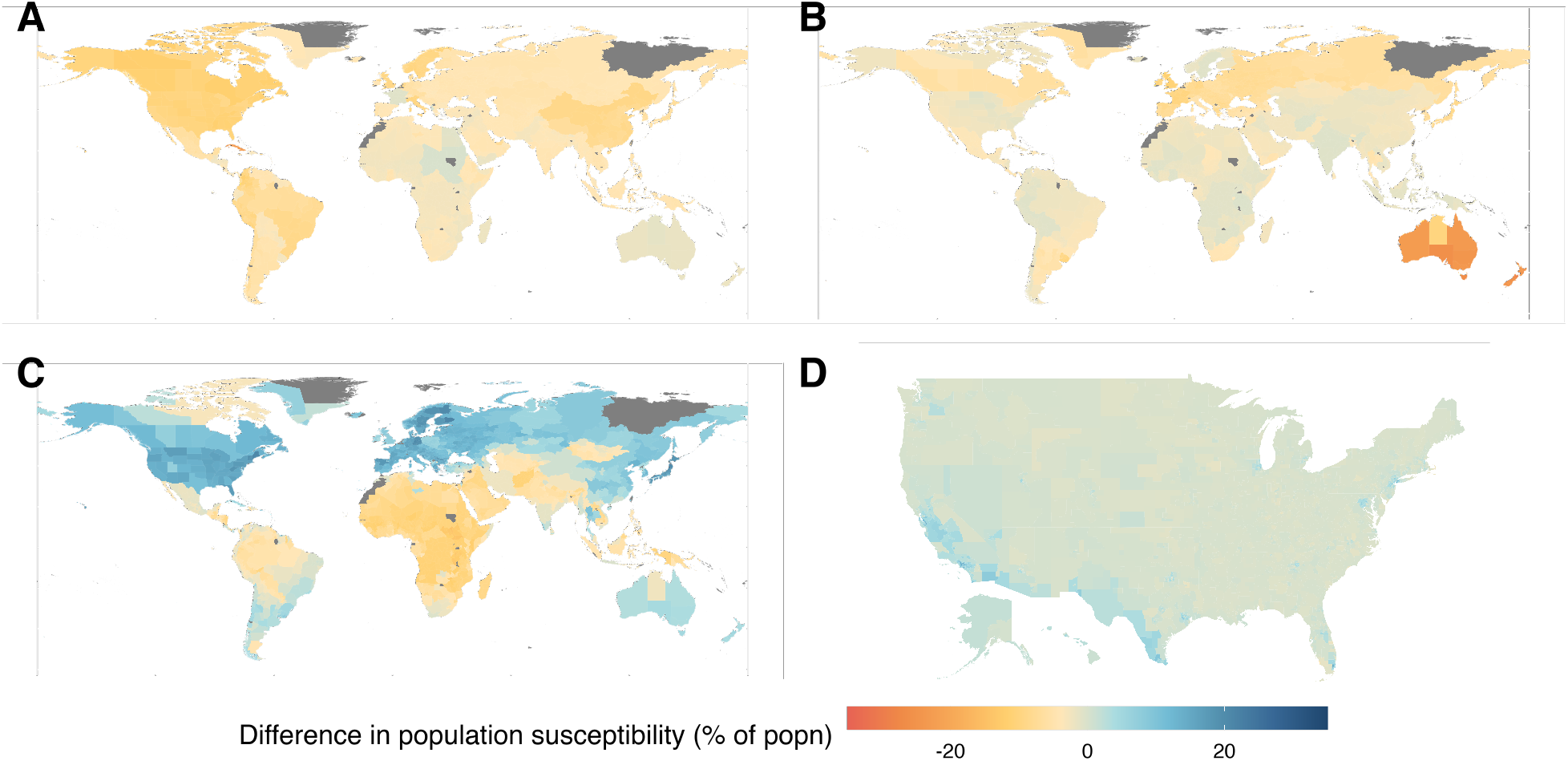
The role of demography and vaccination in shaping the orthopoxvirus susceptibility landscape. Differences in population susceptibility due to four counterfactual scenarios: (A) All countries ceased routine smallpox vaccination in 1984. (B) All countries achieved 100% vaccination coverage before cessation. (C) All administrative regions have the same global average age distribution. (D) There has been no immigration into the U.S. in the last century.

These results further demonstrate that demography is the largest contributing factor to the monkeypox susceptibility landscape and allow us to geographically target these effects nationally and sub-nationally.

The simultaneous cessation scenario (Figure 3A) highlights that later smallpox vaccination cessation would lead to a systematic global decrease in population susceptibility. The effect of later cessation is small in most nations, with some of the largest average decreases of 24% in Cuba and 11% in Canada, countries which stopped routine smallpox vaccination relatively early. The universal vaccination counterfactual (Figure 3B) similarly leads to small decreases in global population susceptibility, with the exception of Australia and New Zealand which had particularly low coverage.

In contrast, the homogeneous demography scenario (Figure 3C) reveals significant changes in the landscape of susceptibility. These changes are greatest in countries with age structures that deviate substantially from the global average: African, southern Asian, and northern South American nations experience an increase in average age under this counterfactual, leading to lower susceptibility. Countries in North America, Europe, northern Asia, and Australia experience decreases in average age, increasing susceptibility substantially. At a finer geographical scale, we find that the north/south gradients observed in India and Brazil (in Figure 1A) are driven by differences in age distributions (Figure S9). Results assuming homogeneous demography within countries are shown in Figure S10.

Lastly, we consider how the immunity landscape would differ in the U.S. if there had been no immigration in the last century, and find notable increases in susceptibility along the U.S./Mexico border and in large urban areas (e.g., Chicago, D.C., NYC, Miami, Houston). The increase in susceptibility arising from the exclusion of immigrants highlights that foreign-born individuals residing in the U.S. have decreased susceptibility to orthopoxvirus infection on average (65% population susceptibility among foreign-born residents versus 73% among U.S.-born residents). This result reinforces the demographic effect we highlight above as foreign-born individuals in the U.S. tend to be older than U.S.-born individuals (45 versus 38, on average), and the residents of high immigration locations in the U.S. tend to have a lower average age.19

## Discussion

Despite the massive success and organizational prowess of the global smallpox eradication campaign, fine-scale vaccination coverage estimates are limited and only focus on the period near eradication four decades ago. Contemporary estimates are needed for public health officials to make informed decisions in the context of increasing orthopoxvirus emergence. We address this gap by creating a comprehensive, high-resolution global map of current susceptibility to orthopoxvirus morbidity, accounting for demographic changes since smallpox was eradicated and possible waning of cross-protective immunity. We illustrate that there is a distribution of substantial global protection against orthopoxvirus diseases. However, we identify significant, previously undocumented, spatial heterogeneity in susceptibility, at national and subnational scales, and demonstrate that this is driven chiefly by differences in demographic change since eradication, and less by differences in historical smallpox vaccination efforts.

The 2022 global monkeypox outbreak has highlighted the immediate relevance of mapping this susceptibility landscape, but our work has broader implications for future risks from all orthopoxviruses. As we get further from smallpox eradication, and the fraction of human populations with any acquired immunity to orthopoxviruses (from vaccination or infection) dwindles, other human-infecting orthopoxviruses stand to benefit from the increasing pool of susceptible individuals (known as ‘competitive release’ in ecological terms).^15^ As the transmission of a number of orthopoxviruses is on the rise, our work quantifies the geographic distribution of vulnerability to this ensemble of emerging viruses. In particular, we convert data on *Vaccinia*-based vaccine distribution and effectiveness against clinical monkeypox and smallpox infection to a global map of susceptibility. However, the effectiveness and durability of *Vaccinia*-based vaccines has not been quantified for all zoonotic orthopoxviruses, so absolute estimates of susceptibility cannot be produced (though for some viruses, like buffalopox, their close genetic relationship to *Vaccinia* makes substantial cross-protection likely). The contribution of our work is in the characterization of the spatial patterns of orthopoxvirus susceptibility, which are driven primarily by vaccination history, not effectiveness, so the relative heterogeneities will be preserved. Thus our mapping shows the landscape of relative risk from any human-infecting orthopoxvirus; and determining the waning rates for all orthopoxviruses to quantify the scale of susceptibility remains a key research frontier.

It is also important to note that our susceptibility estimates primarily quantify vulnerability to symptomatic disease, and the implications for infection or onward transmission are not clear. Smallpox vaccination is expected to reduce risk of infection with monkeypox virus, but quantitative estimates of such sterilizing immunity are limited.^20^ Data on infectiousness of individuals who become infected despite being vaccinated are even more scarce. Here we focus on population-scale susceptibility which is critical for epidemiological risk analyses and public health decision-making, but at the individual level, susceptibility is closer to dichotomous, with unvaccinated individuals generally vulnerable to infection and disease, and vaccinated individuals generally protected. Average susceptibility of different age cohorts may also depend on the unknown proportion of individuals who were vaccinated multiple times pre-eradication, which significantly increases the duration of immunity.^14, 21^ The age-assortative nature of human contact patterns, which can amplify age-specific infection risks, increases the potential impact of this heterogeneity in individual susceptibility.

The process of finding and aggregating country-specific data from frequently incomplete historical records brings inherent uncertainty to the data and our estimates. To capture age-specific vaccination coverages, we leverage scar survey data from each nation, where available. Not all scar surveys were carried out near vaccination cessation, so we assume constant vaccination coverage from the year of a scar survey to cessation of routine vaccination in each country. It is thus possible that decreased coverage near elimination could lead to slightly greater susceptibility in cohorts born in the 1970s. On the other hand, some countries continued some vaccination beyond cessation and eradication (e.g., the DRC had a vaccination rate of 41% in 1981 declining to 4% by 1985^22^) and thus our susceptibility estimates may be conservative for cohorts born near cessation. Due to lack of data, we do not have smallpox vaccination coverage estimates at the admin-1 level. Therefore, we assume that scar surveys are nationally representative and uniform. Where available, we incorporate admin-1 variability in vaccination coverage and cessation into our uncertainty analysis, and find that these assumptions do not significantly reduce confidence in our susceptibility estimates. For a significant number of countries (61%), we were unable to find information on vaccination coverage. We filled this gap with estimates from neighboring countries in 40% of these cases and call on the global health community to assist in filling remaining data gaps. These data limitations underscore the importance of organized reporting and record-keeping of vaccination patterns and disease cases through centralized and modern surveillance systems. Looking forward, modern data collection approaches would be critical to validate and extend our susceptibility estimates. In particular, recent calls for large-scale, routine, and representative serological surveys to characterize population immunity can play a crucial role in our spatiotemporal understanding of the dynamics of pandemic and endemic infections alike.^23^

In this study, we compile the first high-resolution global map of the orthopoxvirus immune landscape using demographic and smallpox vaccination data. Our work reveals the vast global spatial heterogeneity of orthopoxvirus immunity, highlighting the need to consider spatial clustering of susceptible individuals, whether due to age or other factors, and underscoring the importance of fine-scale spatial analysis in light of increased risk of orthopoxvirus emergence and establishment. In response to the 2022 global monkeypox outbreak, we contribute an open database of all our sub-national estimates and uncertainties as an immediate resource for the global health community. We also invite the community to contribute both historical information and future data (e.g., on the distribution of the ACAM2000 and JYNNEOS vaccines) to this living database. The hope remains that the current outbreak will be contained, but in the event that transmission becomes widespread, our monkeypox vulnerability map can inform public health efforts on assessing risks of geographic introductions, allocation of scarce vaccine supplies, and predicting transmission dynamics in concert with data on contact patterns, mobility, and real-time prevalence.

## Methods

Our goal is to generate geographic estimates for population susceptibility to monkeypox at the sub-national level globally. To achieve this, we generate a database of routine smallpox vaccination campaign cessation dates, estimated smallpox vaccination coverage before cessation, and age compositions for each admin-1 region globally. In the U.S., we also incorporate information about country of birth and generate estimates at a finer spatial scale than admin-1. From this database, we estimate the proportion of each population that is expected to have protection from smallpox vaccination and then estimate susceptibility to orthopoxviruses, after accounting for the cross-immunity between smallpox vaccination and smallpox or monkeypox.

### Data

Our database containing information on smallpox vaccination and demography (as detailed below) can be accessed in Tables S1 and S2. In addition to recording the source of each data point, we also graded the quality of evidence for each country.

#### Vaccination cessation

Cessation dates reflect the end of routine smallpox vaccination programs after country-specific smallpox elimination or global smallpox eradication. Global vaccine cessation date information was obtained through a literature review for each country (see dates, sources in Table S1). If a vaccine cessation date was unavailable, the cessation date was assumed to be 1980 based on WHO recommendations to discontinue vaccination that year;^1^ to reflect this assumption, we recorded the lowest data quality grade for this data point (Table S1). Military vaccination after cessation, in the U.S. and other countries, is relevant but was not included in our main estimates, due to an anticipated negligible effect on population susceptibility (details in SI).

#### Vaccination coverage

Vaccination coverage data was obtained, whenever possible, via the most recent scar survey conducted in each country (Table S1). Scar survey information was first obtained in Fenner et al. or the WHO Final Report, then a search of WHO eradication documents and general literature search.^1, 3^ If multiple within-country coverages were given in a scar survey, the population weighted average was taken as the national coverage. If country-specific coverage was not available but coverages for at least two other countries in the region were known, the coverage was determined by averaging known coverages of the closest countries in the region. If age-specific coverage rates were available, the 5-14 age group coverage was used for all individuals of that age range at the time of the scar survey or born between the scar survey and the cessation date, while the ≥15 age group coverage, if available, or the overall coverage, was used for all individuals born 15 years or more prior to the scar survey (see SI). If no age-stratified information was available, the overall population coverage was used, if available. If coverage information could not be obtained, coverage was assumed to be 80% based on the WHO target vaccination coverage for the eradication campaign,^1^ and the lowest data quality grade was recorded (Table S1).

For the U.S., only a national estimate for smallpox vaccination coverage was available.^1^ To capture sub-national spatial heterogeneity in vaccination, we estimated the state-level variation in smallpox vaccination coverage based on polio vaccination (details in SI).

#### Demography

For global estimates, admin-1-level age composition data from the Gridded Population of the World (GPW) dataset^24^ in 5 year age classes from 2010 was re-scaled to 2020 population sizes. For U.S. estimates, we used the Public Use Microdata Sample (PUMS) dataset from the U.S. Census, which provides a sample of individuals representative at the scale of PUMAs, which are geographic areas containing at least 100,000 individuals.^25^ For each individual, the age, location of birth (U.S. state if U.S.-born or country if foreign-born), and current PUMA of residence is known. When aggregating to the population scale, we took into account the survey weights of the PUMS dataset.

### Population susceptibility estimates

To generate orthopoxvirus population susceptibility estimates for each admin-1 level region in all countries, we used the smallpox vaccination cessation dates and reported vaccination coverages from our global database for each country, as well as demographic information.

For global estimates, we first estimated the proportion of a 5-year age group that is expected to be vaccinated with smallpox *Vaccinia* vaccine based on age at cessation and coverage at cessation within a country (Figures S11 - S29). To estimate admin-1-level smallpox susceptibility, we took a population-weighted mean across age-specific vaccination estimates, scaled this value by the estimated effectiveness of smallpox vaccination against monkeypox infection (80 · 7%^8, 10^), and took the complement.

A similar approach was followed for the U.S. estimates, except we estimated the likelihood of a group being vaccinated against smallpox based on age as well as location of birth. For U.S.-born individuals, the coverage in their state of birth and the U.S. cessation date was used. For non-U.S.-born individuals, we used the coverage and cessation date in their country of birth. All estimates are aggregated by current PUMA of residence (accounting for migration away from location of birth).

#### Role of vaccine effectiveness

We examined a higher (less conservative) smallpox vaccine effectiveness of 85%, consistent with estimates from epidemiological analyses of a number of monkeypox outbreaks, as well as a lower effectiveness of 75.6%, factoring in potential waning (estimated based on a linear regression and extrapolation of all available monkeypox vaccine effectiveness data, see SI and Figure S30).

To generalize our susceptibility estimates to other orthopoxviruses, we considered a smallpox vaccine effectiveness of 91 · 1% (an average of the values found during *Variola major* outbreaks) and 74.9% (an average of effectiveness found during *Variola minor* outbreaks, SI and Figure S30).

#### Consistency of estimates with field data

To validate our estimates, we compared our estimated age-specific smallpox vaccination coverage to age-specific smallpox scar survey data^8, 26–28^ and antibody study data^29^ from five countries (details in SI).

### Estimating uncertainty in population susceptibility estimates

To quantify the robustness of our population susceptibility estimates under varying data quality, we characterized the uncertainty in smallpox vaccination cessation dates and coverage levels. We derived uncertainty intervals for coverage and cessation date for each country by placing bounds on plausible values for those inputs and characterizing the part of the interval with the highest likelihood. We then parameterized probability distributions based on these bounds and skew, and performed parametric bootstrapping to estimate population susceptibility with uncertainty. Uncertainty intervals for cessation dates were defined based on known information about each country’s smallpox vaccination campaign. Intervals for vaccination coverage were derived from within-country or regional spatial variability in coverage from scar surveys. For locations without uncertainty information, default uncertainty intervals were assumed (details in SI).

### Estimating the role of demography and vaccination in population susceptibility

To assess the role of each factor in our population susceptibility estimates, we removed heterogeneity in a given demographic or vaccination factor and compared the resulting population susceptibility estimates against the original estimates. The four scenarios we considered are: (1) all countries ceased routine smallpox vaccination in 1984, the last date that any country stopped vaccination; (2) all countries achieved 100% vaccination coverage before cessation, removing heterogeneity in coverage; (3) all administrative regions have the same global average age distribution, removing admin-1 level heterogeneity in age; and (4) there is no immigration into the U.S., meaning foreign-born individuals are excluded from the population susceptibility estimates (details in SI).

## Data Availability

All data and estimates will be available online at https://github.com/bansallab/mpx_landscape.

## Code and data availability

All code to reproduce figures and data to produce our estimates, as well as the estimates are available at https://github.com/bansallab/mpx_landscape.

## Authors’ contributions

JCT and ECR contributed equally: JCT performed the analyses, interpreted the findings, drafted, and edited the manuscript; ECR collected the data, interpreted the findings, drafted, and edited the manuscript. JOL-S interpreted the findings and edited the manuscript. SB conceived and supervised the study, performed analyses, interpreted the findings, and edited the manuscript.

## Acknowledgments

We thank Stephanie Geng for help joining the GPW and GADM datasets for mapping.

## Supplementary Information

### S1 Vaccination coverage heterogeneity

For global estimates, if age-specific coverage rates were available, the 5-14 age group coverage was used for all individuals of that age range at the time of the scar survey or born between the scar survey and the cessation date, while the ≥15 age group coverage, if available, or the overall coverage, was used for all individuals born 15 years or more prior to the scar survey. Country-specific vaccination policies varied in the age of primary smallpox vaccination, with some children vaccinated at 3-4 years of age and others when they entered primary school at 5-6 years of age. By using the 5-14 age group, we ensure that the entire age group is eligible for the primary smallpox vaccination and have more complete coverage data since all members of the age group can be vaccinated.

For the U.S., we estimated spatial heterogeneity in smallpox vaccination coverage (for which data was not available) based on state-level 1995 vaccination rates for 24 month old children with ≥3 doses of polio vaccine. We chose polio vaccination as it was a comparable routine childhood vaccination against a pathogen nearing elimination in the U.S. when the data were collected. We used 1995 data because this was the earliest year the data were provided by the CDC.^30^

### S2 Estimating uncertainty in population susceptibility estimates

In order to address uncertainty inherent in aggregating historical or incomplete data, we used a parametric bootstrapping approach. We placed bounds on plausible values for the cessation date and vaccination coverage for each country to represent the uncertainty at that level. We estimated a probable distribution of values within these bounds and drew 5000 samples for vaccination coverage and cessation date for each country to generate the range of plausible outcomes under this level of uncertainty. With each pair of independently sampled cessation dates and scar survey coverages, we re-estimated population susceptibility, generating 5000 estimates, from which we took the mean and standard deviation for each admin-1 region.

We estimated bounds on cessation date values depending on whether the cessation date was known for a given country. If the cessation date was known, the lower bound of uncertainty was that date and the higher bound was one year after the known cessation date with a right-skewed distribution to reflect higher likelihood on the known cessation date and the possibility of a brief delay in enacting the policy change during which vaccination continued. If no cessation date was known, a default interval of +/− 2 years was used with a symmetric distribution with a mode at the 1980 WHO-recommended cessation date. For some countries, WHO documents reported a higher bound at which point vaccination no longer occurred, but the lower bound was unknown. In these cases, the lower bound is assumed to be 1970 as this was among the earliest known cessation dates globally, and a uniform distribution was used to reflect the lack of a known or estimated cessation date during this period.^31^ For more information on the type of data available, the estimates and ranges used, and sources by country, see Table S1.

Depending on where our highest certainty fell in the cessation date range, we sampled possible cessation dates from a `beta-binomial` distribution (for the symmetric, left-skewed, and right-skewed cases) or a discrete uniform (for cases where we lacked confidence in a most likely value). For the symmetric case, we used *d*_*L*_ + `Beta-Binomial`(*n, α* = 1000, *β* = 1000); for the right-skewed case, we used *d*_*L*_ + Beta-Binomial(*n, α* = 0 · 4, *β* = 1 · 5); and for the left-skewed case, we used *d*_*L*_ + `Beta-Binomial`(*n, α* = 1 5, *β* = 0 4). In all cases, *n* was parameterized by the difference between the upper bound date and lower bound date, and *d*_*L*_ was the lower bound date.

We estimated uncertainty on vaccination coverage values, where possible, using known spatial heterogeneity in coverage levels from admin-1 regions within a country or from neighboring countries. In these cases, the bounds and the shape of the distribution of plausible values were represented by the empirical distribution of within-country or regional coverage values, whether it be symmetric, left-skewed or right-skewed, with a mean defined by the average of the available values. In cases where a country’s known scar survey did not include admin-1 coverages, a default interval of +/− 5% from the overall coverage was used with a symmetric distribution with the mean at the overall value. In cases where the default coverage of 80% was used, a uniform distribution with default interval of +/− 5% was used. For more information on the type of data available, the estimates and ranges used, and sources by country, see Table S1.

Depending on where our highest certainty fell in the coverage range, we sampled possible coverage values from a transformed beta distribution (for the symmetric, left-skewed or right-skewed cases) or a uniform distribution. We parameterized and transformed the beta distribution as such: *c*_*low*_ +(*c*_*high*_ − *c*_*low*_)`Beta`(*a, b*), with *b* = 2 and *a* = 2 · (*c*_*mean*_ − *c*_*low*_)*/*(*c*_*high*_ − *c*_*mean*_), and *c*_*low*_ and *c*_*high*_ as the lower and higher bounds of the plausible coverages for a country, and *c*_*mean*_ as the mean of the available coverages.

### S3 Role of natural immunity

We considered potential natural immunity arising from natural smallpox infections prior to eradication and monkeypox outbreaks in endemic countries. Even in endemic areas, monkeypox outbreaks are typically small and self-limiting.^1^ While, monkeypox incidence has recently increased in many endemic areas^33, 34^ larger outbreaks have cumulatively affected very small percentages of the population (e.g., 0 013% of the DRC population) and are therefore likely to be negligible in susceptibility calculations, especially in regions with high vaccination coverage.^8, 35, 36, 38^ However, monkeypox cases in endemic areas may be underestimated due to lack of diagnostics and testing, clinical similarity to other illnesses, and underedeveloped or underutilized disease surveillance systems.^39^

Likewise, we expect the contribution of natural smallpox immunity to the monkeypox susceptibility landscape to be insignificant. In countries with endemic smallpox after 1969, cumulative incidence was extremely low (rarely exceeding 0 · 2% of a country’s population by 1980) and evidence of prior infection was concentrated in adults, who are now 65 or older and often compose a smaller proportion of the age distribution of the affected countries (Figures S4, S5, S9). Overall, the proportion of people naturally infected with either smallpox or monkeypox is markedly outweighed by the relatively high levels of coverage resulting from mass vaccination campaigns that confer much greater levels of population immunity.

As such, we assumed that natural immunity for smallpox and monkeypox was negligible. Even if smallpox incidence was 1.5 times greater than reported figures, we see negligible (< 0 · 5%) differences in susceptibility in countries with endemic spread after 1969 (Figure S6).

#### Vaccine effectiveness: Waning and cross protection

In Figure S30, we present a review of data on *Vaccinia*-based smallpox vaccine effectiveness against monkeypox. The data from monkeypox outbreaks estimate effectiveness as *≈* 85% for outbreaks in the 1980s,^5–7^ and 80 · 7% − 80 · 9% during outbreaks in the 2000s.^8, 10^

In Figure S2, we illustrate changes in our population susceptibility estimates given two other vaccine effectiveness scenarios (to the one considered in the main analysis of effectiveness = 80 · 7%). First, we consider the scenario of 85% to reflect evidence from monkeypox outbreaks in the 1980s^5–7^ (Figure S30).

Second, we consider the scenario of waning vaccine effectiveness. We note that multiple studies have found elevated antibody and memory B cell responses persist over 50 years after vaccination and, although T cells wane after 15 to 20 years, T cell levels remain over ten times that of unvaccinated individuals for decades.^14, 21, 46, 47^ Epidemiological estimates of waning vaccine effectiveness post-eradication are not available, however. To investigate the impact of potential waning in vaccine effectiveness, we consider the scenario of a lower vaccine effectiveness of 75·6%. To arrive at this estimate, we perform a linear regression on all available monkeypox outbreak-derived vaccine effectiveness estimates (Figure S30). We assume a linear relationship for waning vaccine effectiveness due to the limited availability of data, and we warn caution against considering this estimate as epidemiologically accurate.

In Figure S30, we also review the evidence available on *Vaccinia*-based smallpox vaccine effectiveness against smallpox, which remains important given the clinical similarity between monkeypox and *Variola minor* and the potential of *Variola major* as a bioterror threat. The data from *Variola major* outbreaks have a weighted average effectiveness of 91 · 1%,^48–50^ while for *Variola minor* outbreaks the weighted average effectiveness is 74 · 9%.^51, 52^ Our susceptibility estimates for these scenarios can be found in Figure S3.

### S4 Checking consistency to field data

To validate our population immunity estimates, we make comparisons to smallpox vaccination take data.

First, we compare our age-specific estimates of the likelihood of being smallpox vaccinated against recent (i.e., 2003 onward) age-specific data on protection against smallpox from five countries. In particular, we use scar survey data from a census in Guinea Bissau in 2005 with a sample size of 1,751;26 from a census in the Democratic Republic of the Congo in 2006 with a sample size of 4,676;8 from scar survey data of a cross-section of a rural population in the Brazilian state of Minas Gerais in 2012 with a sample size of 240;27 from a scar survey of a population of farm workers and their household members in Colombia in 2016 with a sample size of 134;28 and from an antibody study from routine laboratory samples from Italy in 2003 with a sample size of 523.29 For each of these cases, we compare the population immunity of each age group as estimated by our model to empirical estimates. For comparison to the scar survey data, we estimate the expected smallpox vaccination coverage of each age group at the time of the scar survey relative to vaccination cessation in the given country (e.g., for a 2003 scar survey and a 1971 cessation with 70% vaccination coverage in 0-15 year olds (yo) and 90% coverage in 15+ yo at cessation, the 2003 age-specific vaccination coverage estimates would be: 1-31 yo = 0%, 32-47 yo = 70%, 48+ yo = 90%). For comparison to the antibody study data, we generate expected age-specific smallpox vaccination coverage at the time of the study (as described above) and scale this estimate by the vaccine effectiveness estimate of 80 ·7% as a proxy for serological protection.

In Figure S7, we show that most age-specific model estimates are consistent with age-specific empirical estimates. The estimates for Brazil and Colombia are the least consistent, though both studies have small sample sizes and are in clustered, rural populations.

### S5 Military vaccination after 1980

In estimating orthopoxvirus susceptibility in the U.S., we do not include military vaccination policies due to the small proportion of individuals vaccinated relative to the U.S. population as a whole. Although the

U.S. ended routine smallpox vaccination in 1972, military personnel were routinely vaccinated until 1990. In response to the potential threat of smallpox as a biological weapon, the U.S. military restarted its smallpox vaccination program and has continued vaccinating military personnel since the early 2000s. Between December 2002 and November 2017, approximately 2.08 million military personnel and 39,500 civilian healthcare workers received smallpox vaccinations.^57, 58^ The majority of military personnel vaccinated during this period (71%) were too young to have been vaccinated previously, so are likely not included in our estimates of U.S. population susceptibility. However, the total number of military vaccinees is less than 1 · 5% of the U.S. population between the ages of 18 and 64, and less than 0 · 7% of the general U.S. population,^59^ so incorporating these vaccinations would likely result in minimal changes to our population susceptibility estimates. Other countries may have conducted military-specific vaccination campaigns as well, although these efforts also are unlikely to contribute significantly to population susceptibility estimates.

### S6 Age distribution data and mapping

Age distributions procured from Gridded Population of the World (GPW) data^24^ date back to 2010, when some countries had different admin-1-level jurisdictions than today. As a result, to map these old admin-1 divisions to more recent ones documented in the Database of Global Administrative Areas (GADM), we had to merge some GPW admin-1 regions into one GADM admin-1 region, or split other GPW admin-1 regions amongst multiple GADM admin-1 regions.

Maps are created using the Database of Global Administrative Areas (GADM) dataset, version 4.0.^61^

## S7 Supplementary figures

**Figure S1:**
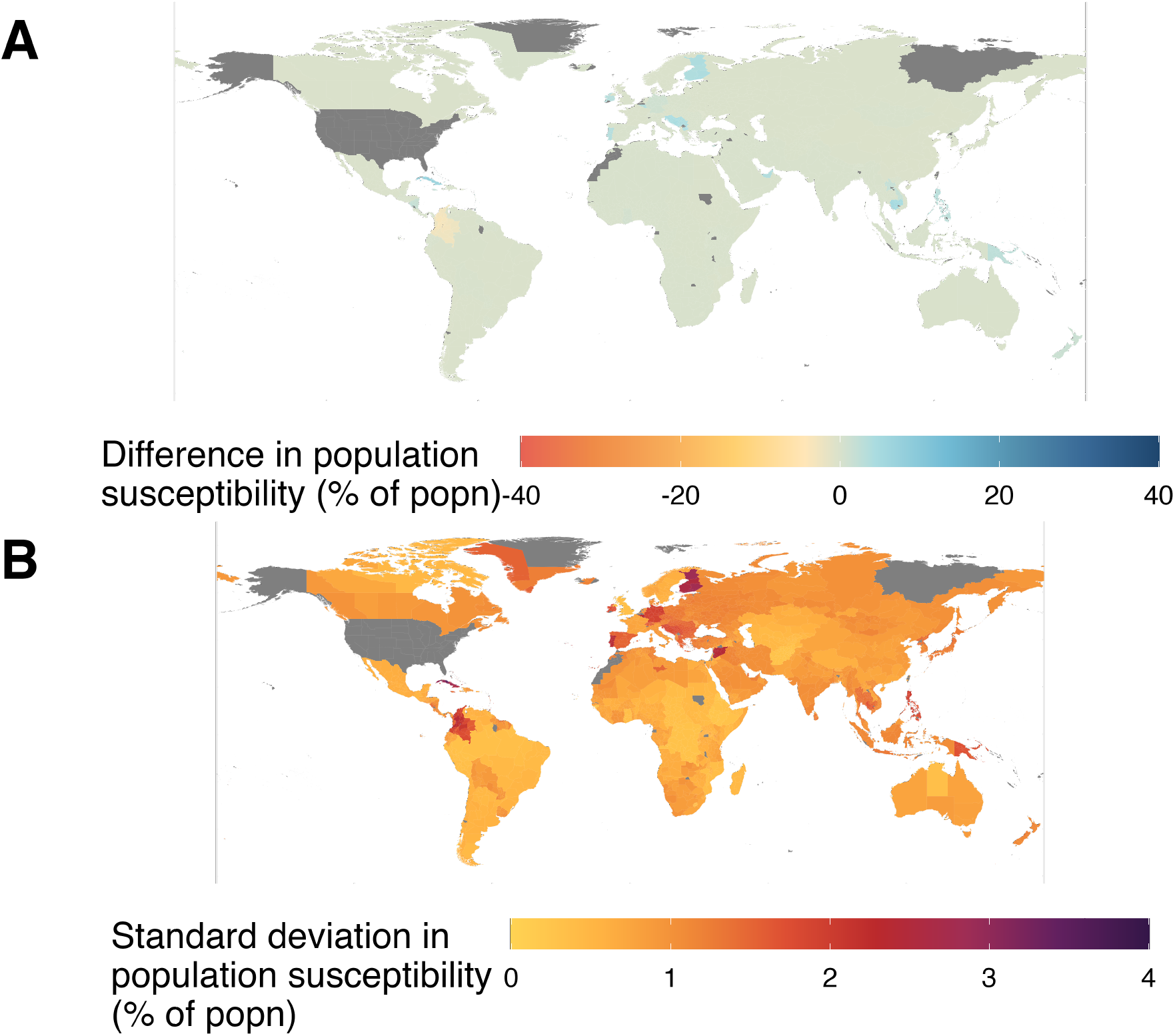
Uncertainty analysis shows results are qualitatively (and quantitatively) robust. (A) Mean difference between original and simulated estimates is greatest in Cuba, Cambodia, and Slovenia. (B) Standard deviation of simulated estimates is greatest in Cuba, Luxembourg, and Finland. Results are from 5000 samples each of vaccination coverage and cessation date distributions.

**Figure S2:**
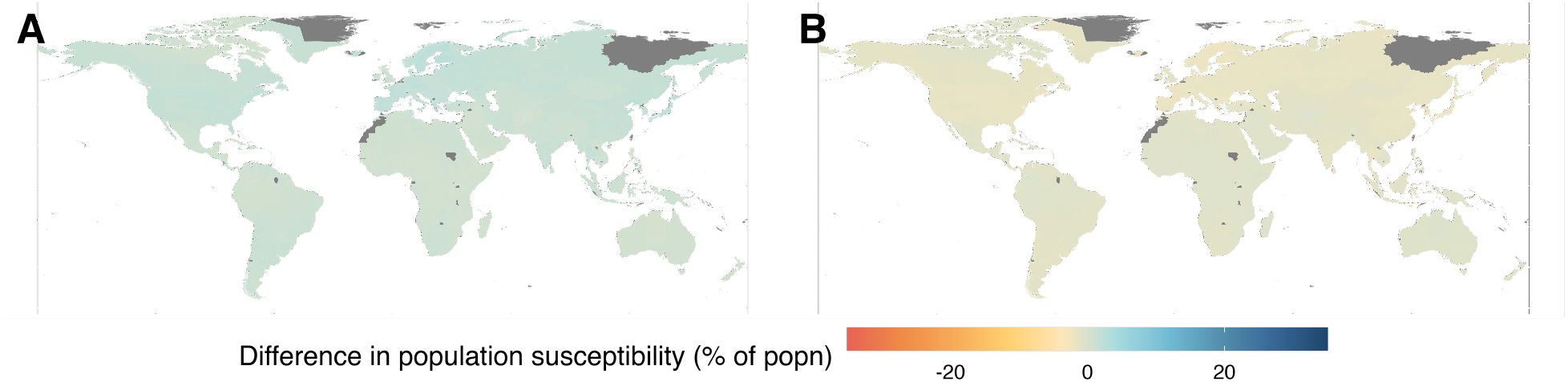
Sensitivity analysis of smallpox vaccine effectiveness against monkeypox infection. (A) 75 · 6% vaccine effectiveness: differences range from 0 27 to 2 7 percentage points of susceptibility from Figure 1A. (B) 85% vaccine effectiveness: differences range from 2 3 to 0 22 percentage points of susceptibility from Figure 1A.

**Figure S3:**
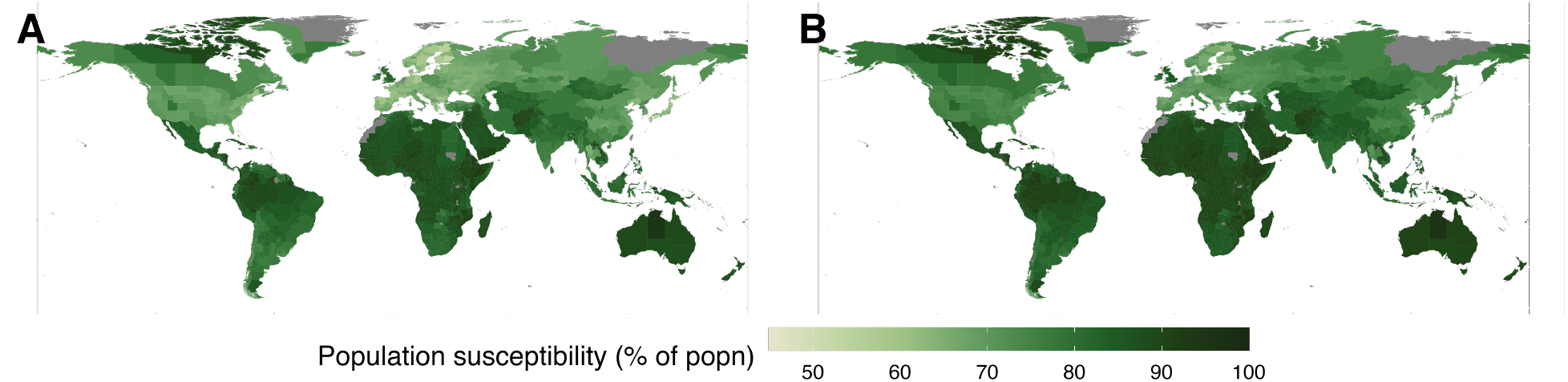
Susceptibility landscape for smallpox infection. (A) *Variola major*, 91 · 1% vaccine effectiveness. Differences from Figure 1A (not shown) range from − 5 · 5 to − 0 · 55 percentage points. (B) *Variola minor*, 74 · 9% vaccine effectiveness. Differences from Figure 1A (not shown) range from 0 · 31 to 3 · 1 percentage points.

https://github.com/bansallab/mpx_landscape/TableS1.csv

Table S1: This table contains raw data on cessation dates, vaccination coverage, estimates of uncertainty intervals, and sources for our estimates.

https://github.com/bansallab/mpx_landscape/TableS2.csv

Table S2: This table contains our estimated admin-1-level 5-year age group population sizes based on 2010 age composition and 2020 total population data from the Gridded Population of the World database.^24^

**Figure S4:**
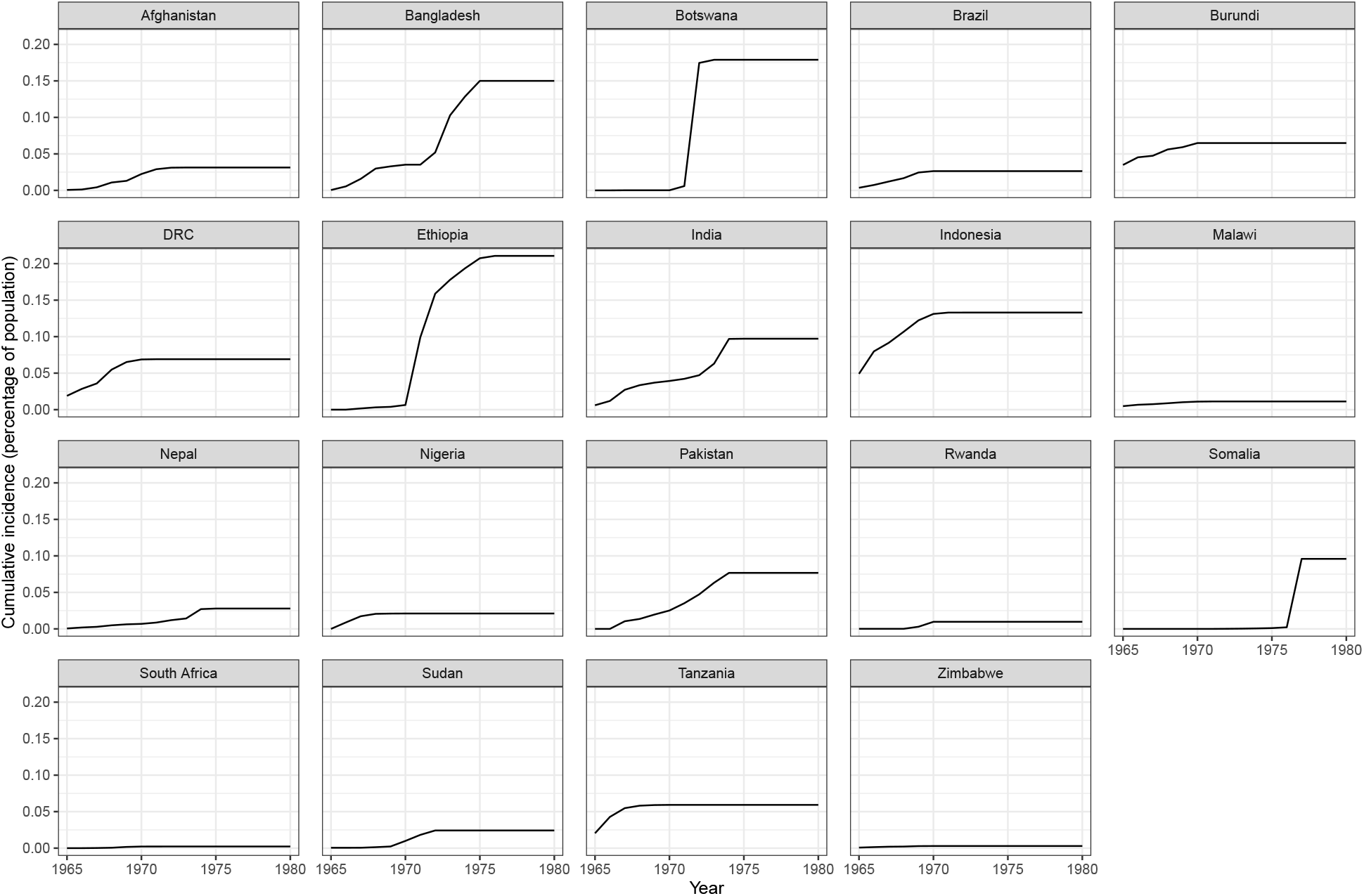
Cumulative incidence in countries with endemic smallpox spread in 1970 or later. Excludes Iran and Iraq due to lack of data. DRC = Democratic Republic of the Congo.

**Figure S5:**
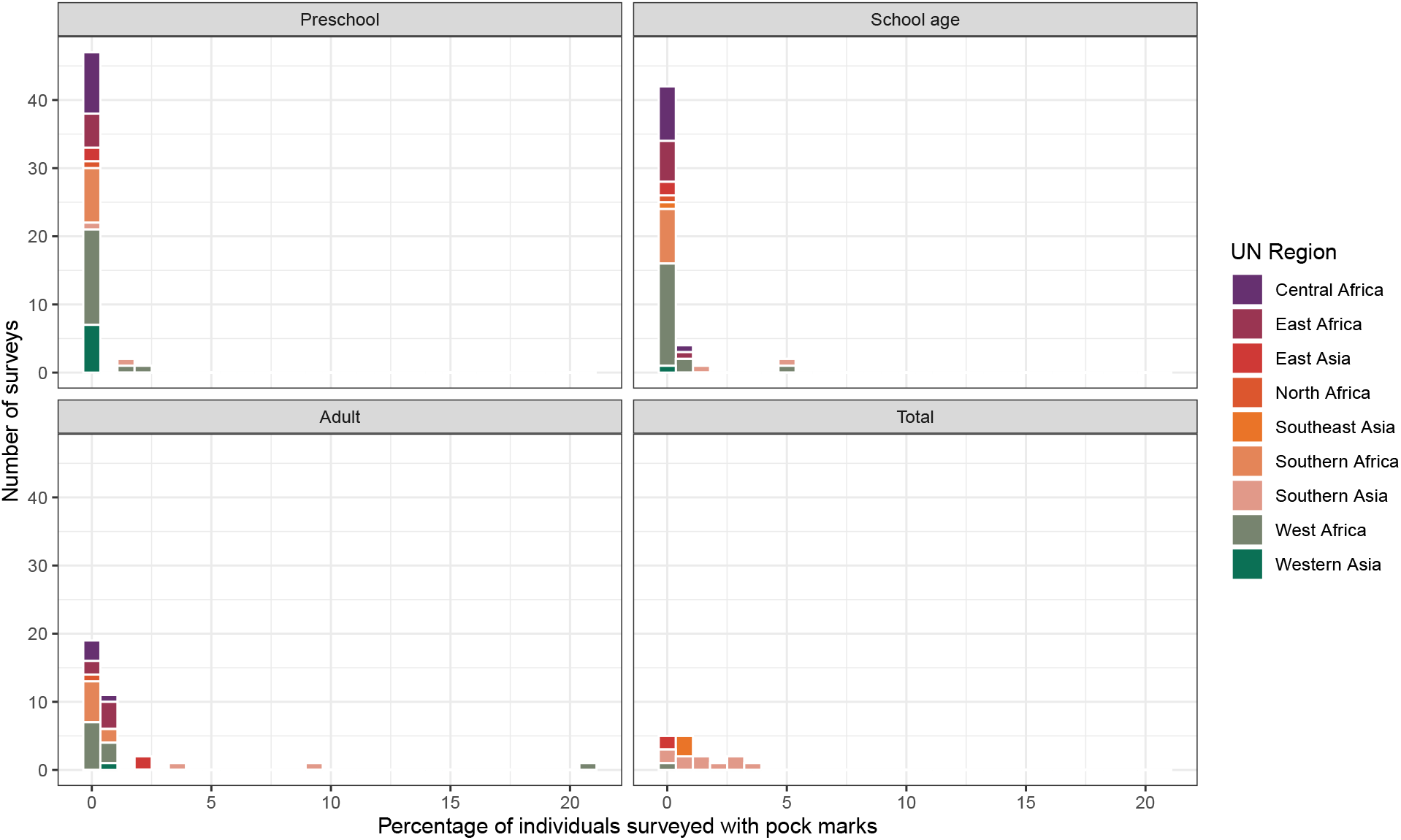
Pockmark survey data indicate few children were naturally infected with smallpox in the 1960s and 70s. Facial pockmark surveys were used to assess the progress of smallpox eradication campaigns, as individuals who had natural smallpox infection generally presented with facial scarring. Here, we use these survey results to demonstrate the low percentages of preschool and school-aged children with signs of natural smallpox infection in the years preceding smallpox eradication. In only 11% of surveys did the percentage of individuals with pockmarks exceed 1% and only in adults did the percentage of pockmarked individuals ever exceed 5%. Adults, who were ≥ 15 years old at the time of these surveys, would be over 65 years old in 2022. Total percentages are provided when an age-specific breakdown of individuals with pockmarks was not available.

**Figure S6:**
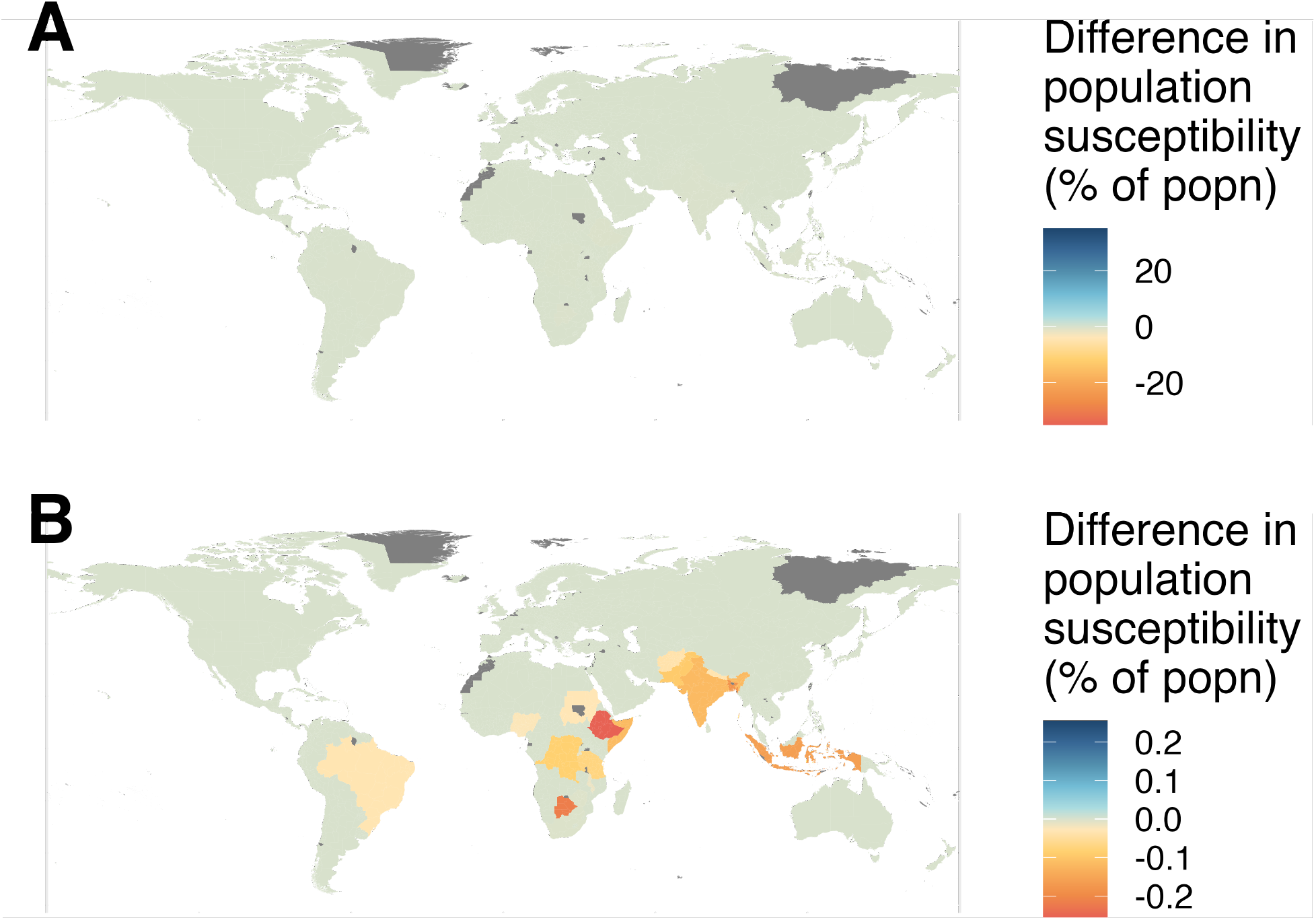
Inclusion of natural immunity has little effect on protection estimates. Assuming smallpox incidence was 1.5 times greater than reported figures and that infections only occurred in unvaccinated individuals, we see negligible (< 0 · 5%) differences in susceptibility in countries with endemic spread after 1969. Panel A uses the same bounds as other figures showing differences from original estimates, while Panel B shows just how small these differences are by narrowing the difference bounds. Excludes Iran and Iraq due to lack of data.

**Figure S7:**
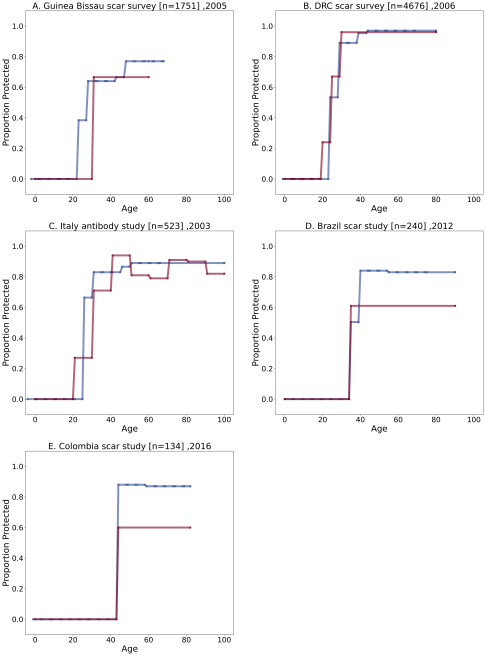
Comparison of our population susceptibility estimates to field data. In panels (A)-(E), we compare the smallpox population immunity (vaccination or vaccination*serological protection) of each age group as estimated by our model (blue) to empirical estimates (red). The varying age ranges for the empirical data reflect the age groups defined by each study. Our model estimates are at 5-year age groups.

**Figure S8:**
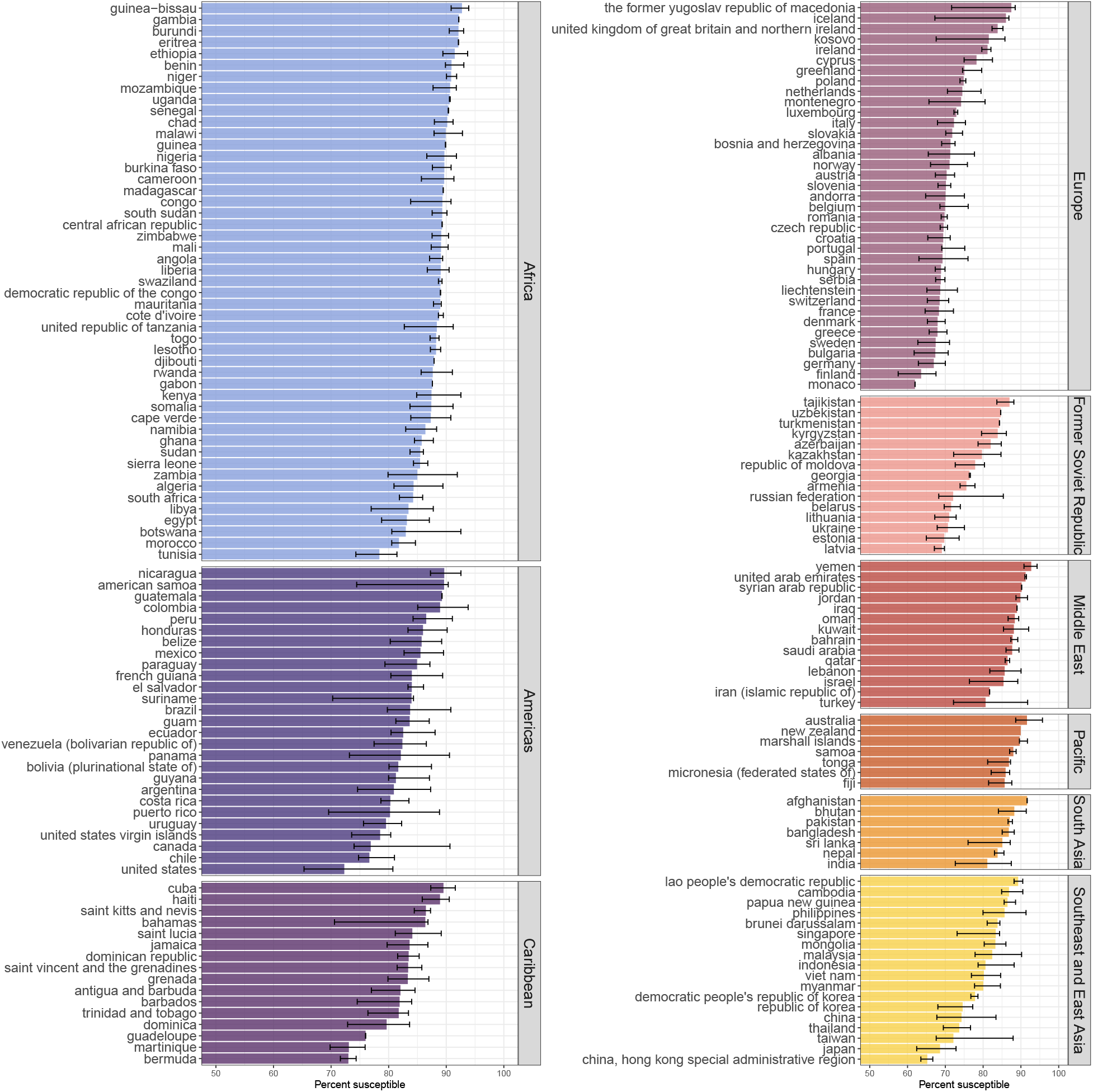
Estimates of population susceptibility by country and region. Error bars show range of estimated susceptibility across admin-1 regions in each country shown in Figure 1A.

**Figure S9:**
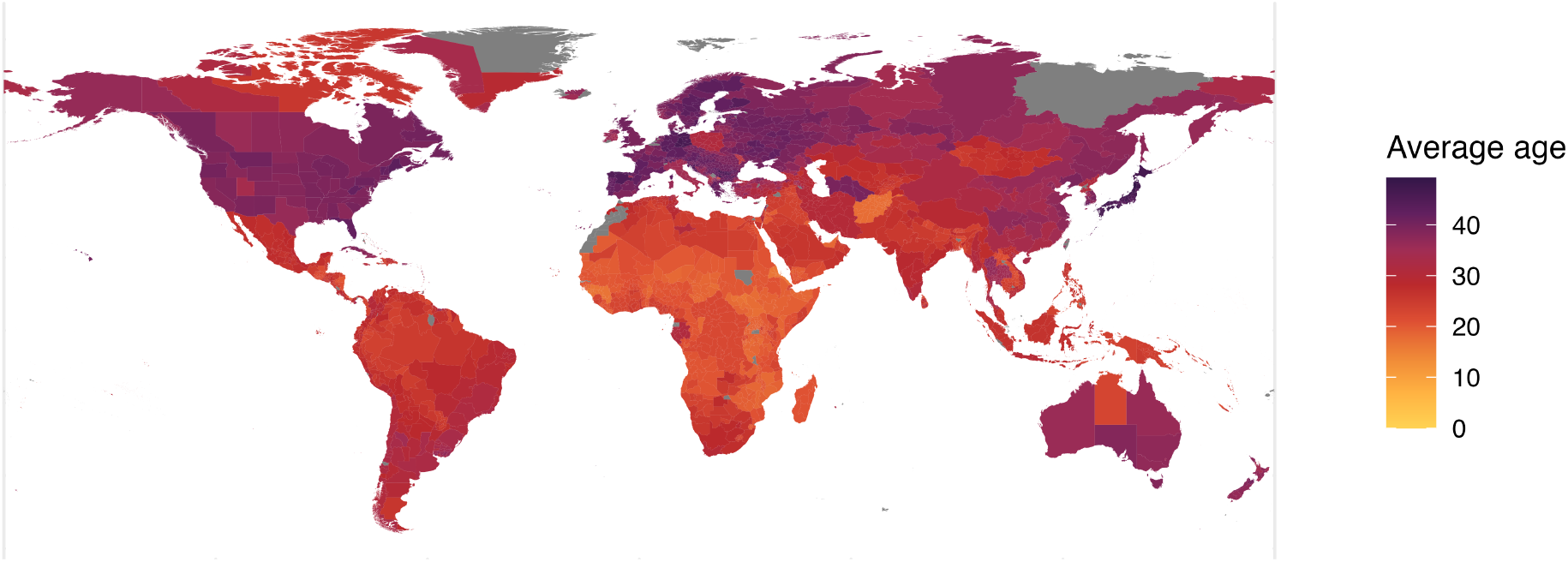
Average age in each admin-1 unit. Average global age is 29 · 6.

**Figure S10:**
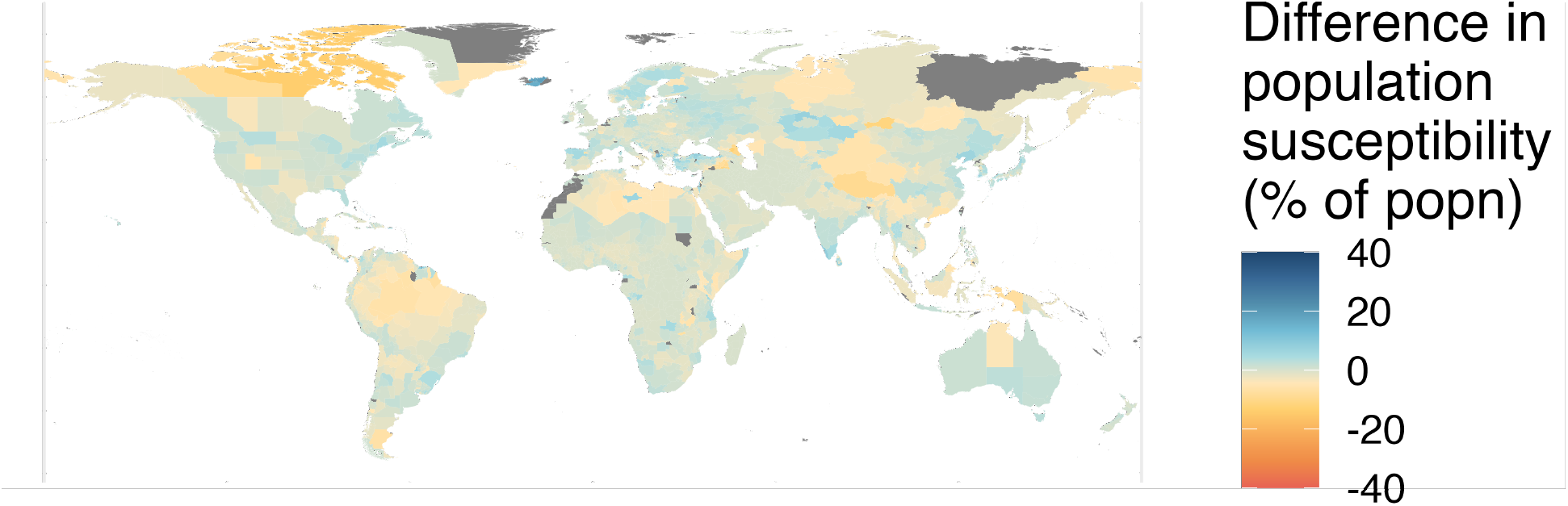
Scenario analysis to identify role of demography in population immunity estimates. In this scenario, we consider homogeneous national demography (i.e., all administrative regions within a country share the the same age distribution as the national average).

**Figure S11:**
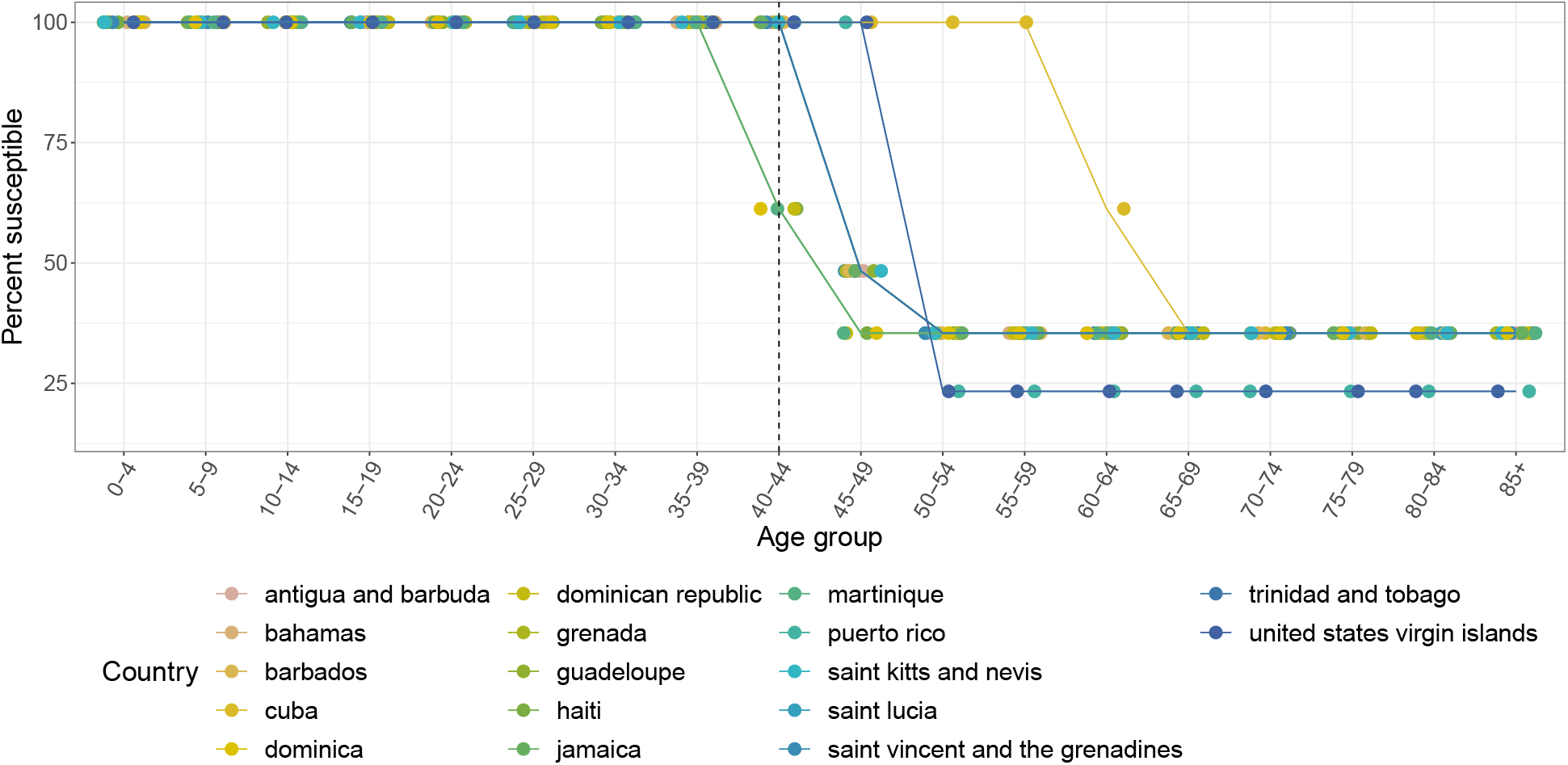
Country-specific monkeypox susceptibility profiles for countries in the Caribbean. Susceptibility is calculated as 1 − (*ϵ* · `proportion vaccinated`) for each 5-year age group, where *ϵ* = 80 · 7% is the smallpox vaccine effectiveness against monkeypox. Dashed line indicates last age group in which some individuals were born before global smallpox eradication (1980). Points are jittered horizontally for visual aid.

**Figure S12:**
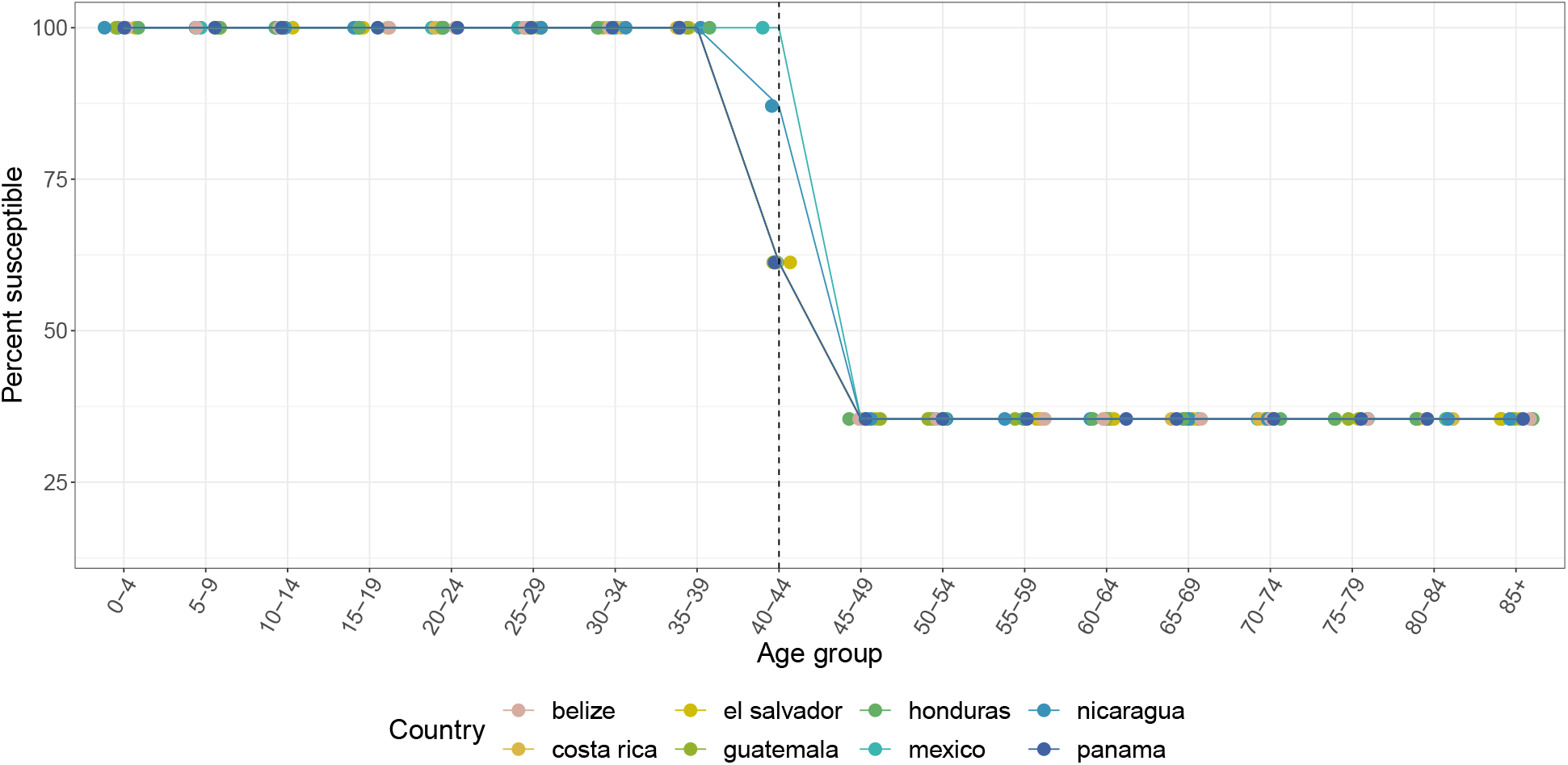
Country-specific monkeypox susceptibility profiles for countries in Central America. Susceptibility is calculated as 1 − (*ϵ* · `proportion vaccinated`) for each 5-year age group, where *ϵ* = 80 · 7% is the smallpox vaccine effectiveness against monkeypox. Dashed line indicates last age group in which some individuals were born before global smallpox eradication (1980). Points are jittered horizontally for visual aid.

**Figure S13:**
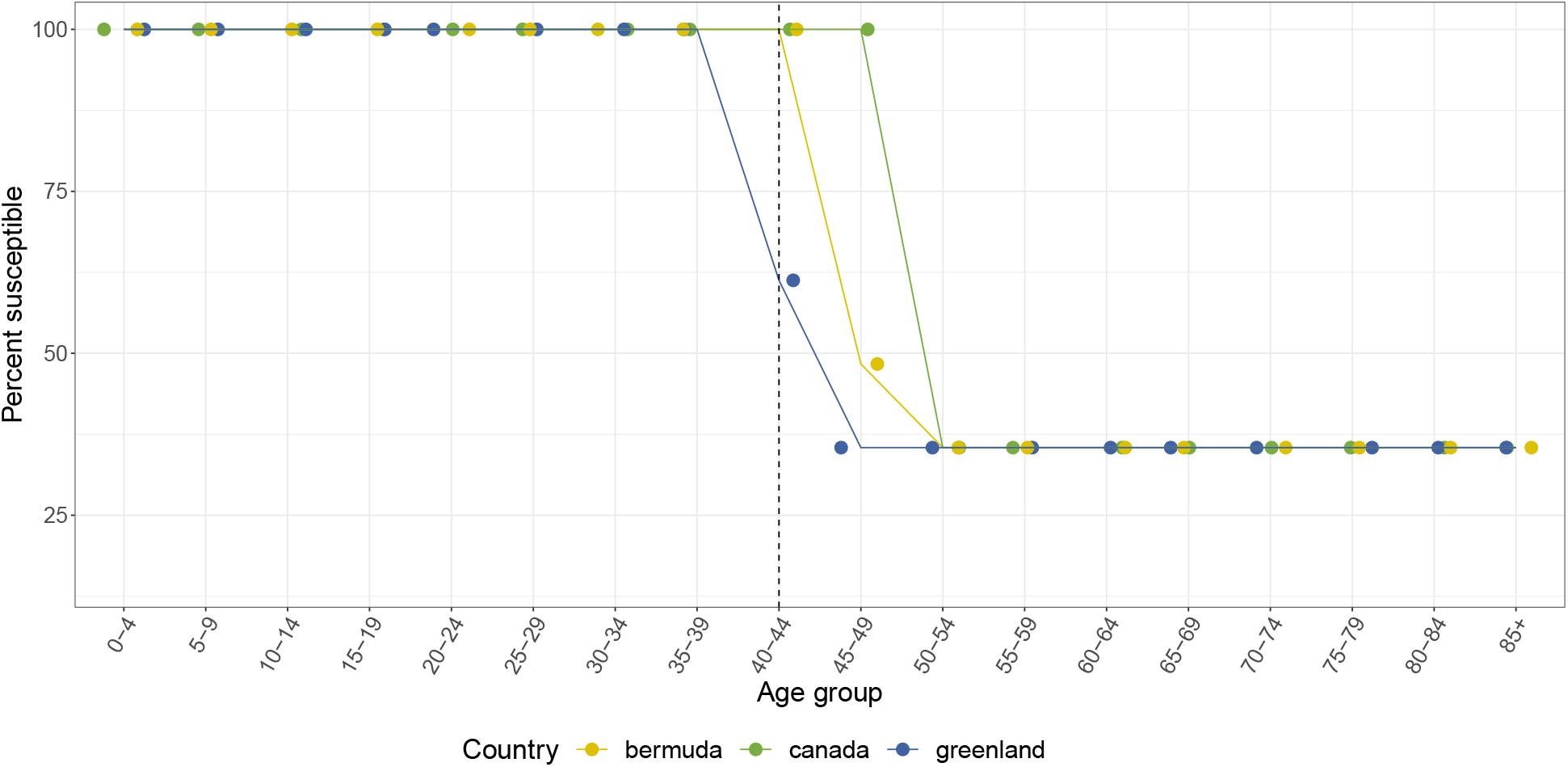
Country-specific monkeypox susceptibility profiles for countries in North America. Susceptibility is calculated as 1 − (*ϵ* · `proportion vaccinated`) for each 5-year age group, where *ϵ* = 80 · 7% is the smallpox vaccine effectiveness against monkeypox. Dashed line indicates last age group in which some individuals were born before global smallpox eradication (1980). Points are jittered horizontally for visual aid.

**Figure S14:**
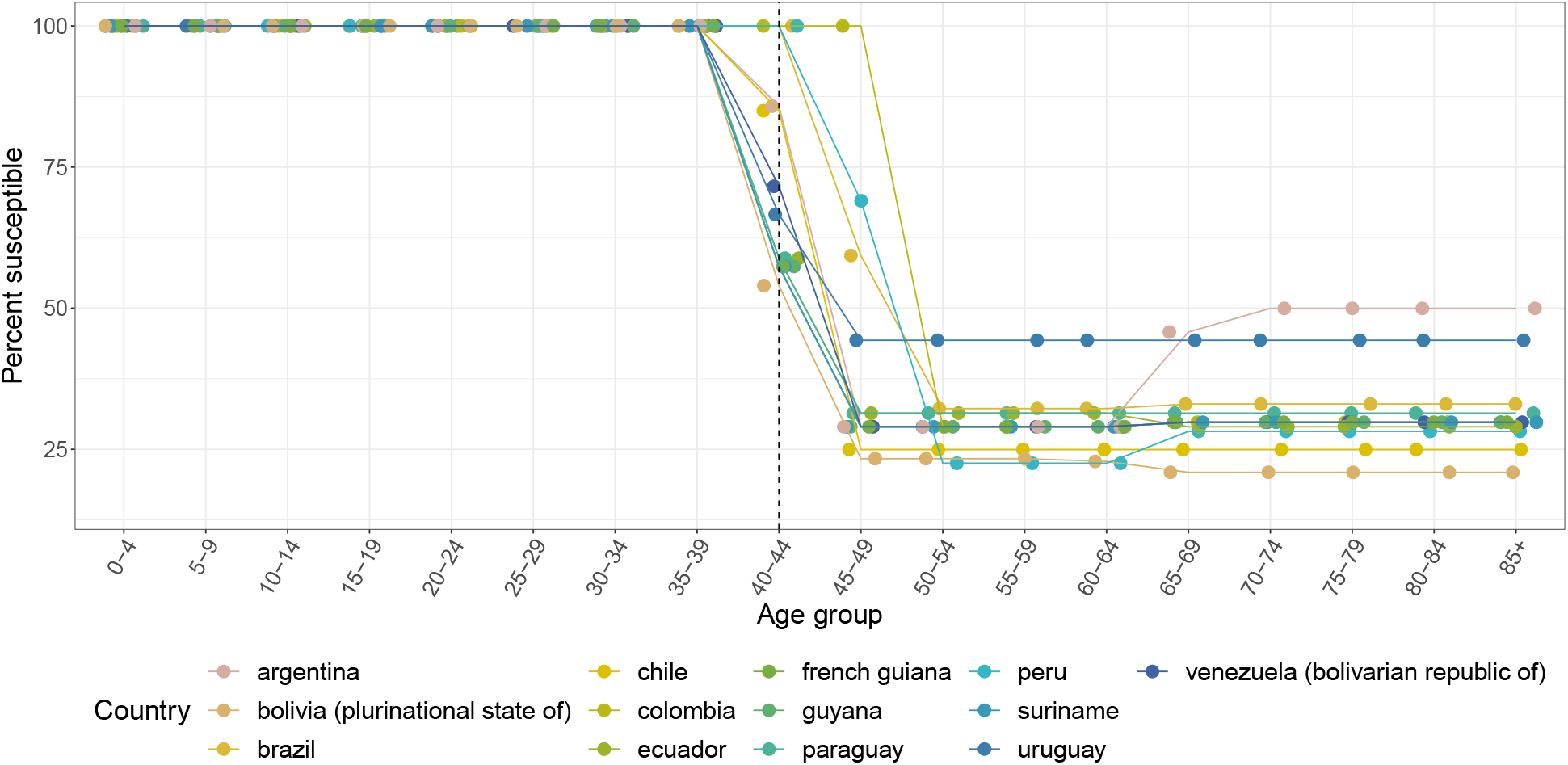
Country-specific monkeypox susceptibility profiles for countries in South America. Susceptibility is calculated as 1 − (*ϵ* · `proportion vaccinated`) for each 5-year age group, where *ϵ* = 80 · 7% is the smallpox vaccine effectiveness against monkeypox. Dashed line indicates last age group in which some individuals were born before global smallpox eradication (1980). Points are jittered horizontally for visual aid.

**Figure S15:**
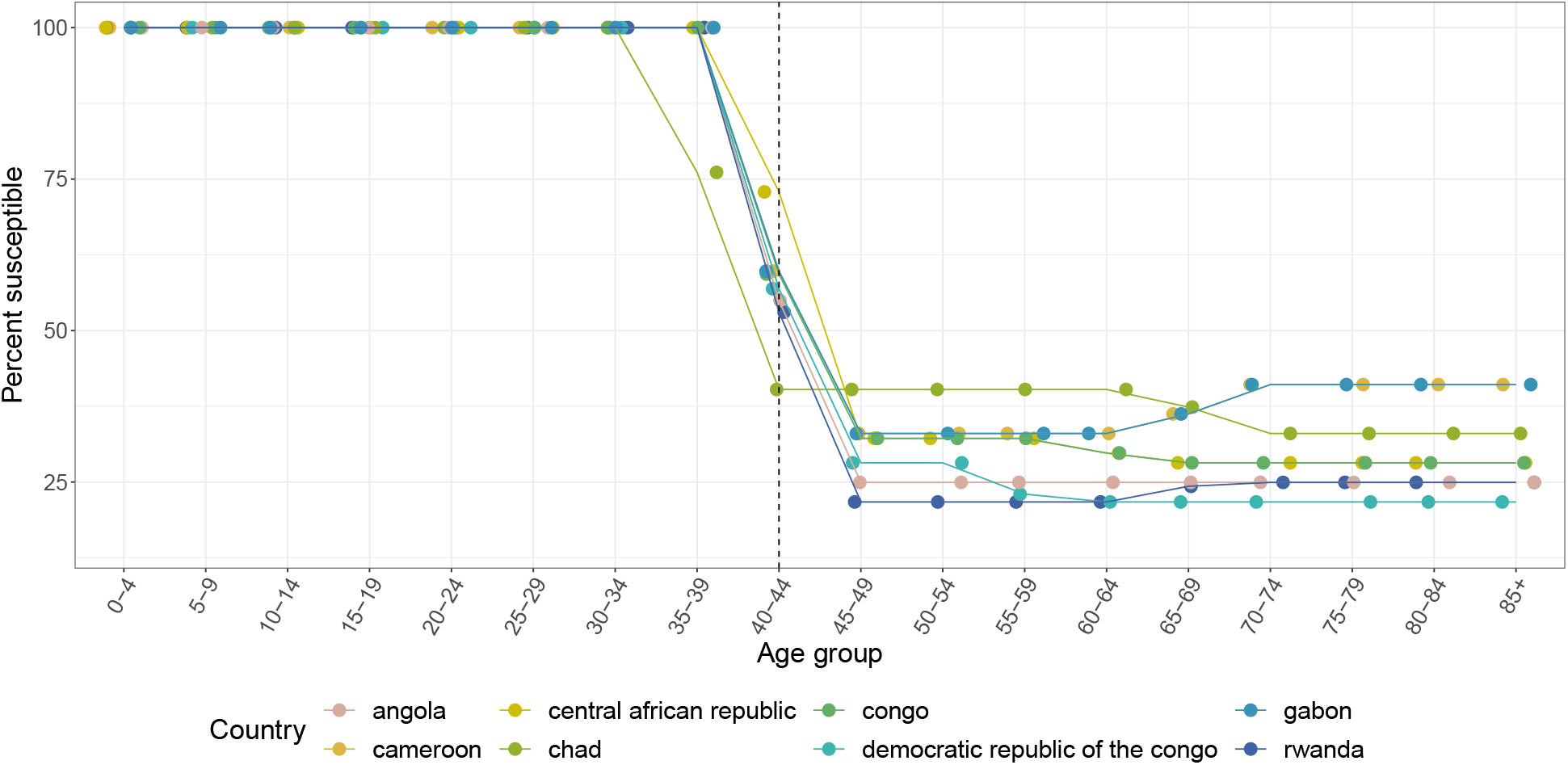
Country-specific monkeypox susceptibility profiles for countries in Central Africa. Susceptibility is calculated as 1 − (*ϵ* · `proportion vaccinated`) for each 5-year age group, where *ϵ* = 80 · 7% is the smallpox vaccine effectiveness against monkeypox. Dashed line indicates last age group in which some individuals were born before global smallpox eradication (1980). Points are jittered horizontally for visual aid.

**Figure S16:**
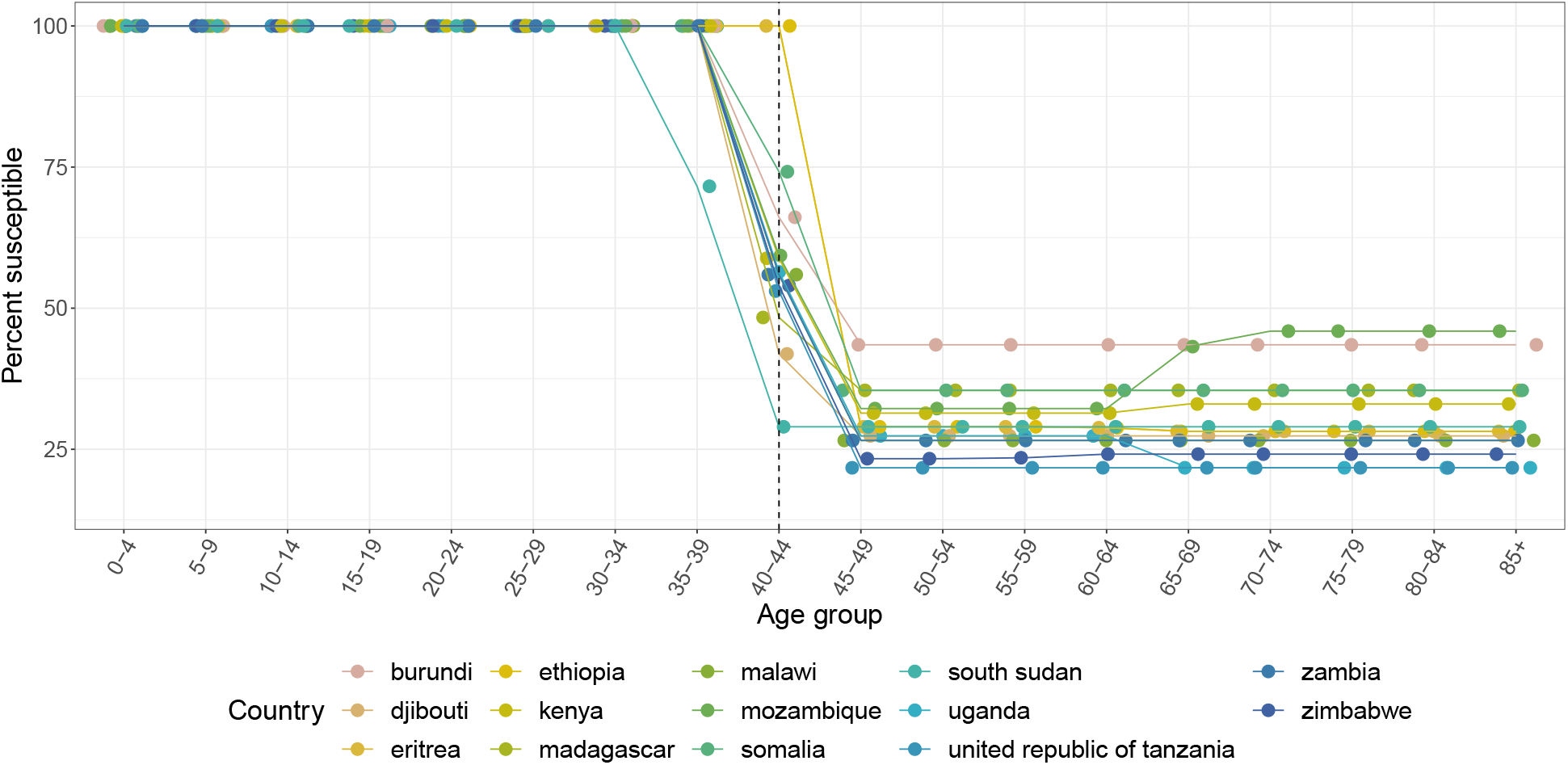
Country-specific monkeypox susceptibility profiles for countries in East Africa. Susceptibility is calculated as 1 − (*ϵ* · `proportion vaccinated`) for each 5-year age group, where *ϵ* = 80 · 7% is the smallpox vaccine effectiveness against monkeypox. Dashed line indicates last age group in which some individuals were born before global smallpox eradication (1980). Points are jittered horizontally for visual aid.

**Figure S17:**
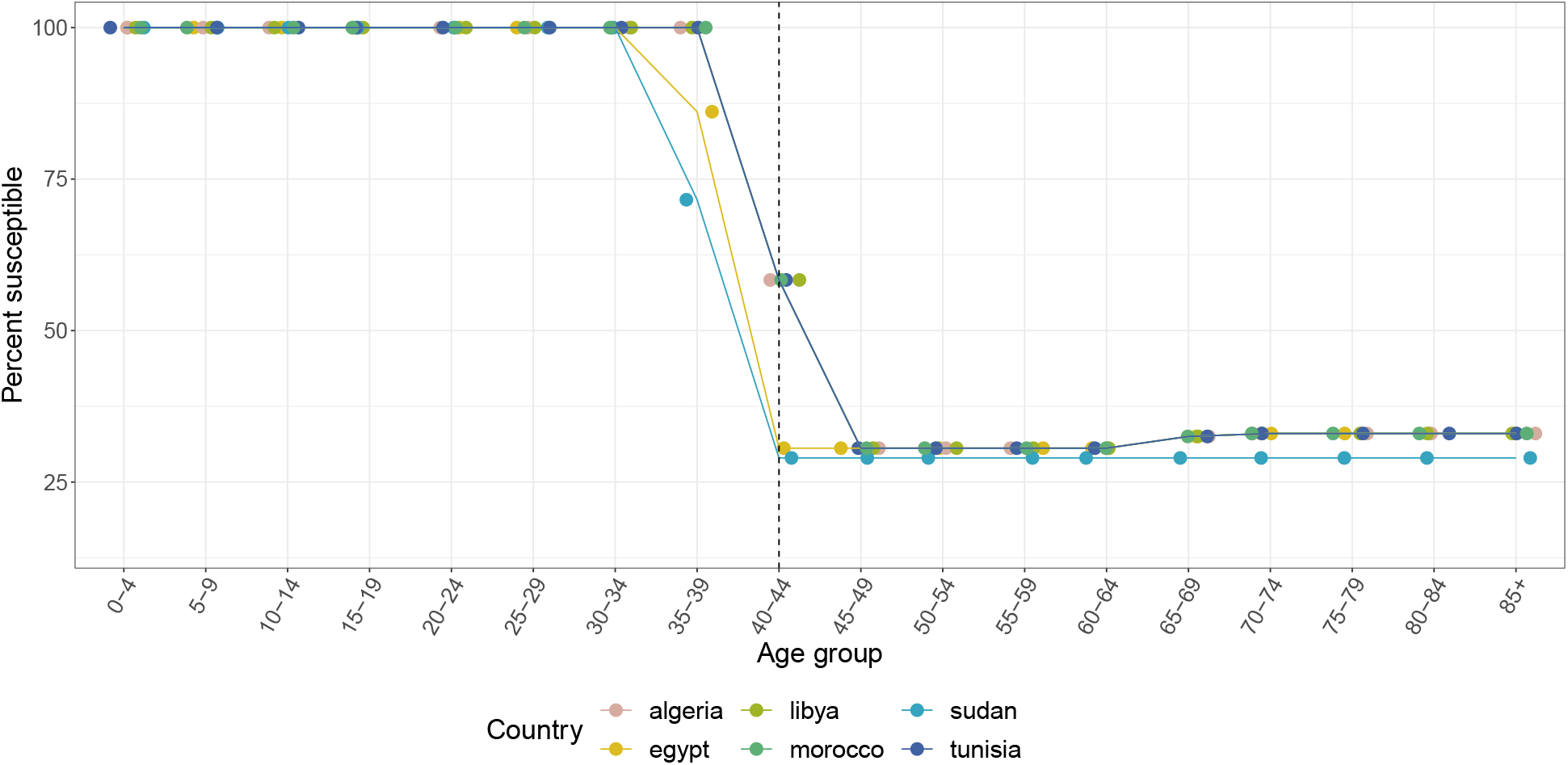
Country-specific monkeypox susceptibility profiles for countries in North Africa. Susceptibility is calculated as 1 − (*ϵ* · `proportion vaccinated`) for each 5-year age group, where *ϵ* = 80 · 7% is the smallpox vaccine effectiveness against monkeypox. Dashed line indicates last age group in which some individuals were born before global smallpox eradication (1980). Points are jittered horizontally for visual aid.

**Figure S18:**
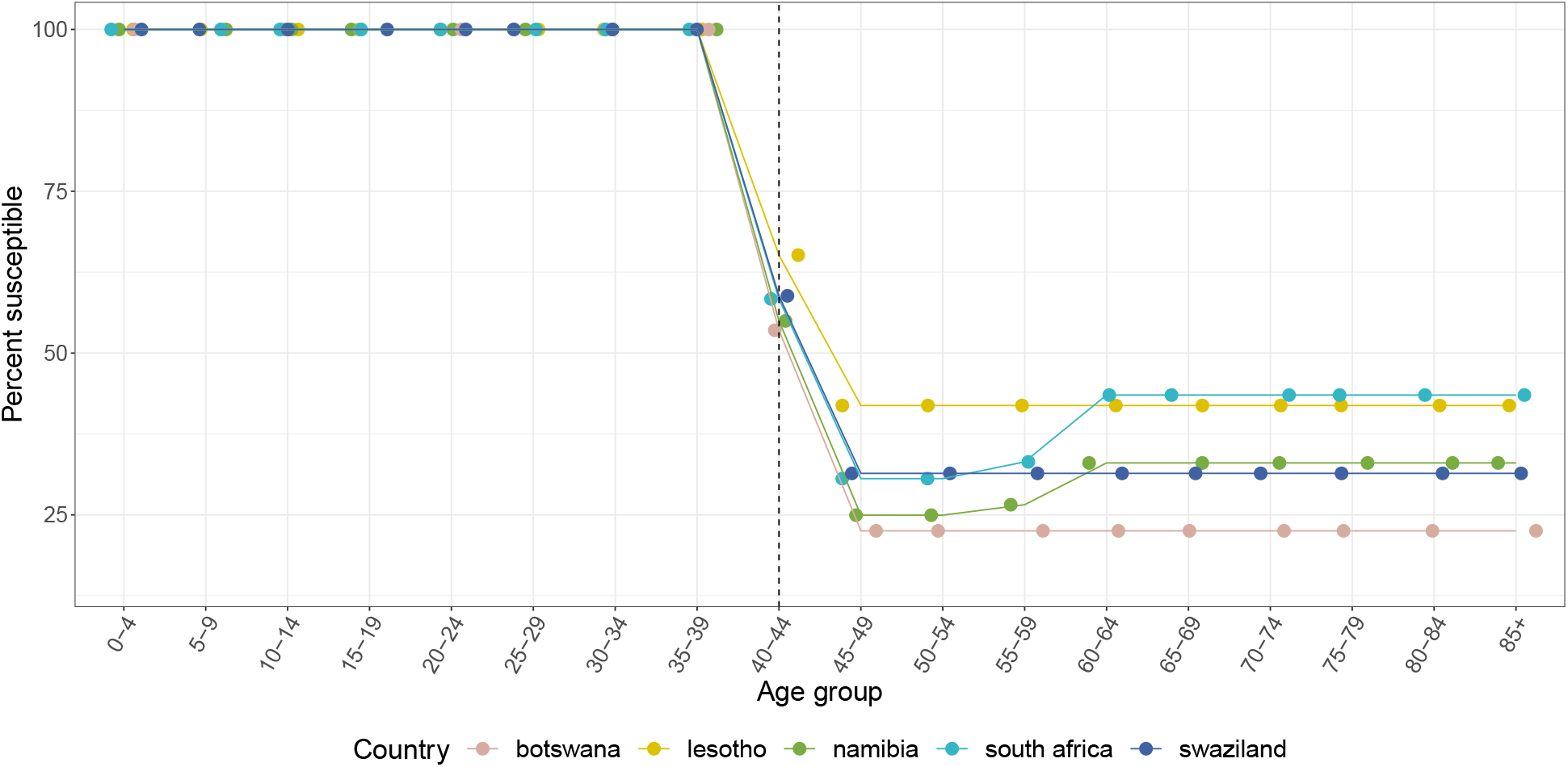
Country-specific monkeypox susceptibility profiles for countries in Southern Africa. Susceptibility is calculated as 1 − (*ϵ* · `proportion vaccinated`) for each 5-year age group, where *ϵ* = 80 · 7% is the smallpox vaccine effectiveness against monkeypox. Dashed line indicates last age group in which some individuals were born before global smallpox eradication (1980). Points are jittered horizontally for visual aid.

**Figure S19:**
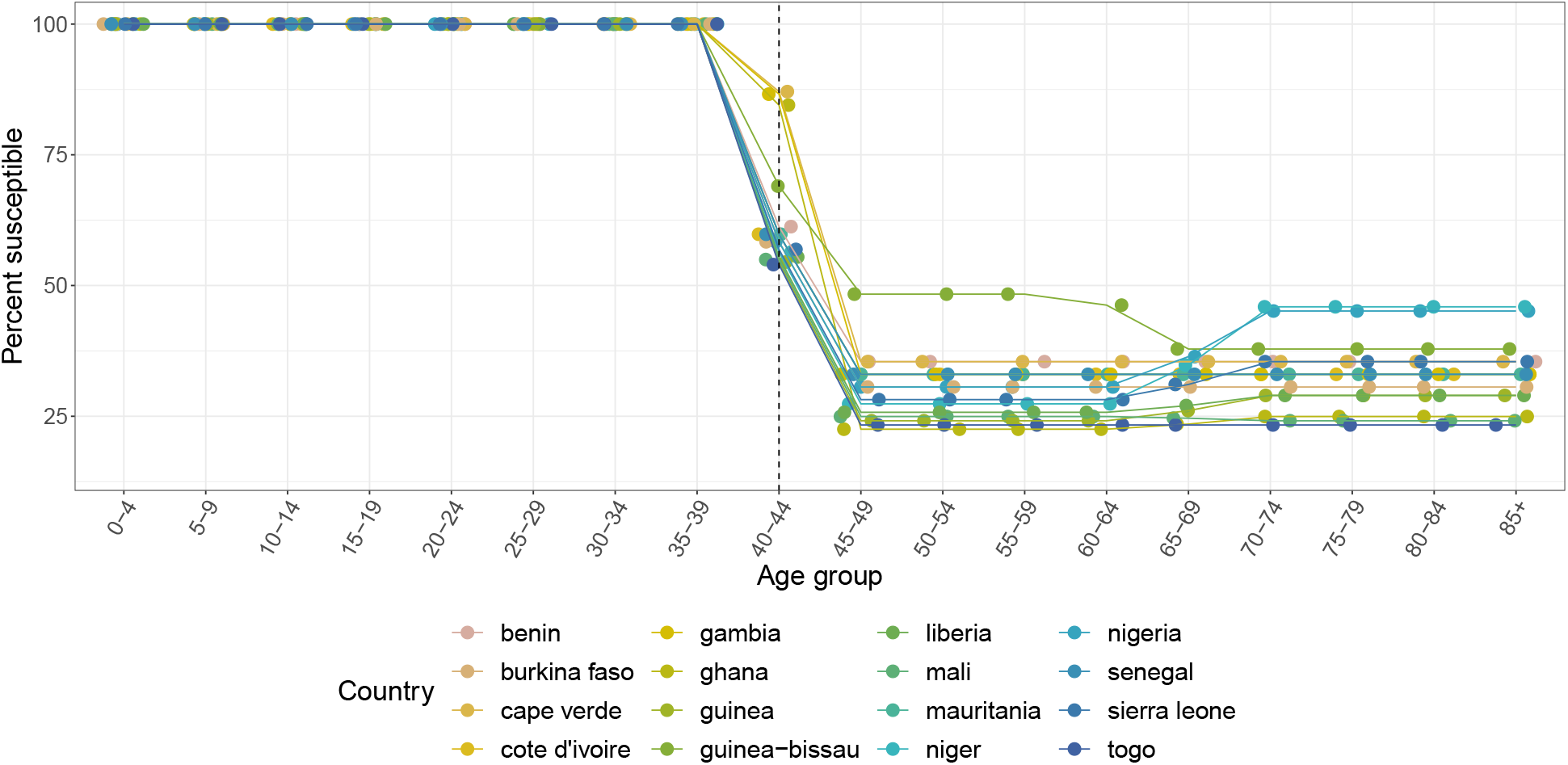
Country-specific monkeypox susceptibility profiles for countries in West Africa. Susceptibility is calculated as 1 − (*ϵ* · `proportion vaccinated`) for each 5-year age group, where *ϵ* = 80 · 7% is the smallpox vaccine effectiveness against monkeypox. Dashed line indicates last age group in which some individuals were born before global smallpox eradication (1980). Points are jittered horizontally for visual aid.

**Figure S20:**
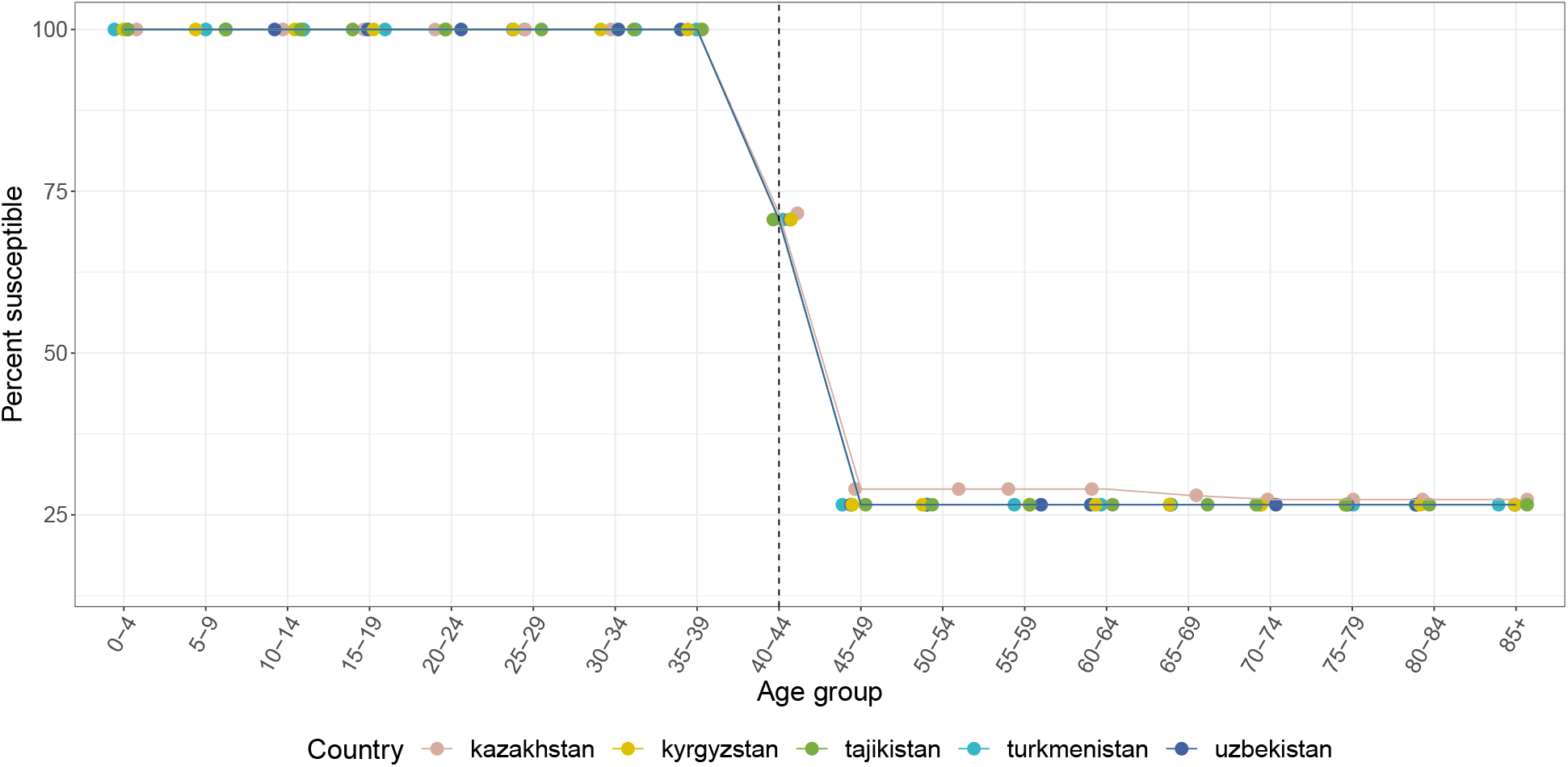
Country-specific monkeypox susceptibility profiles for countries in Central Asia. Susceptibility is calculated as 1 − (*ϵ* · `proportion vaccinated`) for each 5-year age group, where *ϵ* = 80 · 7% is the smallpox vaccine effectiveness against monkeypox. Dashed line indicates last age group in which some individuals were born before global smallpox eradication (1980). Points are jittered horizontally for visual aid.

**Figure S21:**
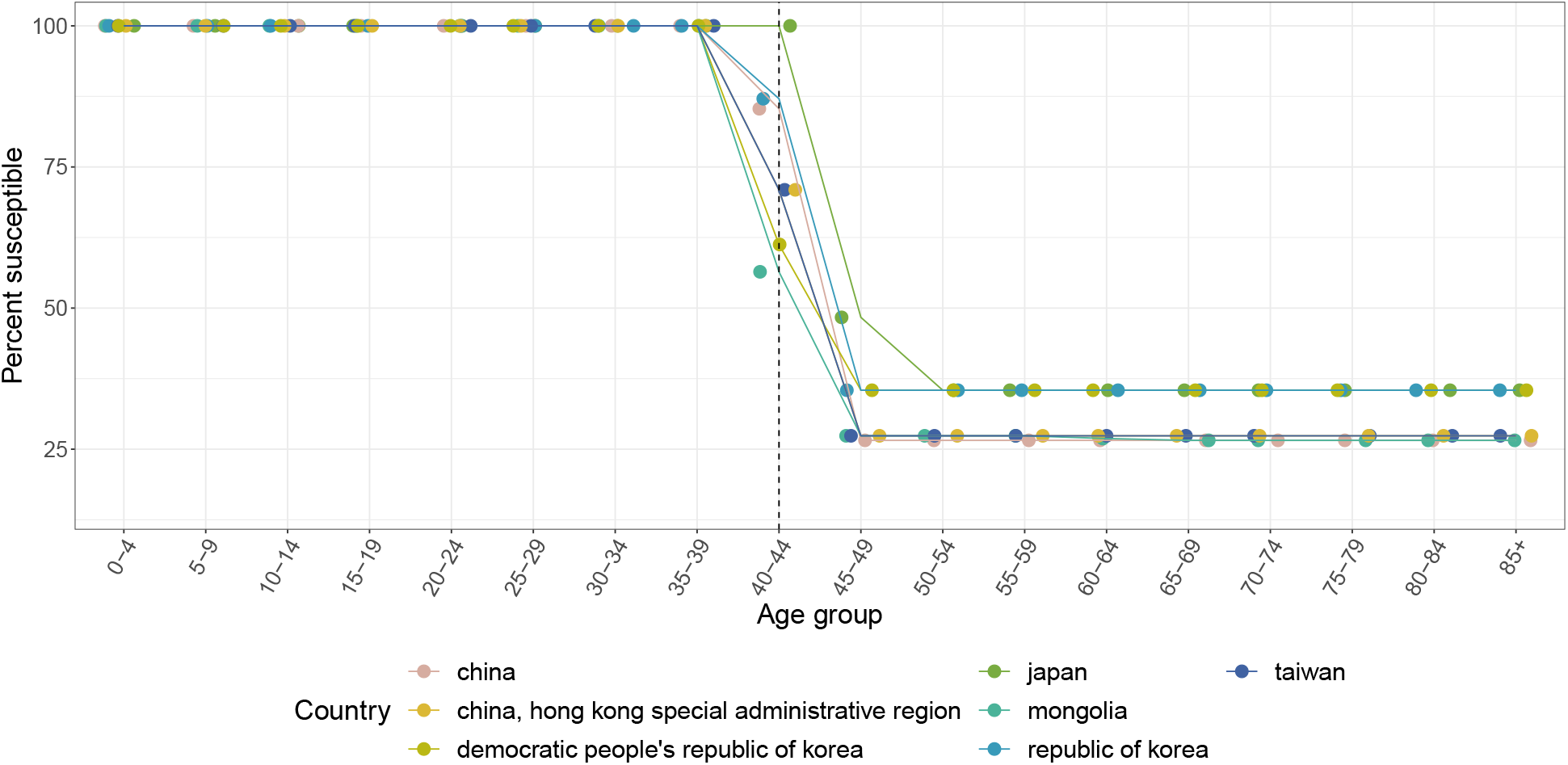
Country-specific monkeypox susceptibility profiles for countries in East Asia. Susceptibility is calculated as 1 − (*ϵ* · `proportion vaccinated`) for each 5-year age group, where *ϵ* = 80 · 7% is the smallpox vaccine effectiveness against monkeypox. Dashed line indicates last age group in which some individuals were born before global smallpox eradication (1980). Points are jittered horizontally for visual aid.

**Figure S22:**
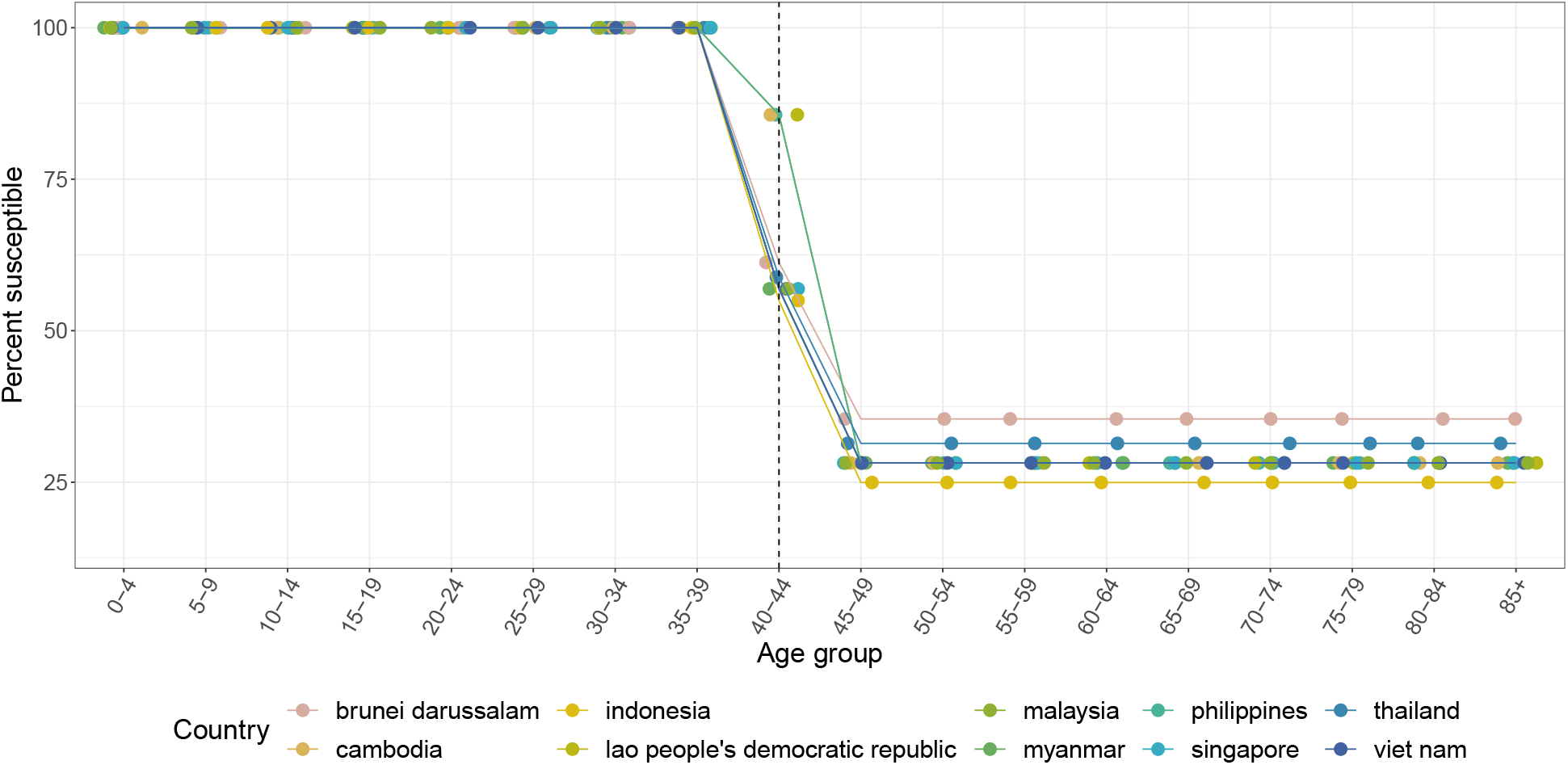
Country-specific monkeypox susceptibility profiles for countries in Southeast Asia. Susceptibility is calculated as 1 − (*ϵ* · `proportion vaccinated`) for each 5-year age group, where *ϵ* = 80 · 7% is the smallpox vaccine effectiveness against monkeypox. Dashed line indicates last age group in which some individuals were born before global smallpox eradication (1980). Points are jittered horizontally for visual aid.

**Figure S23:**
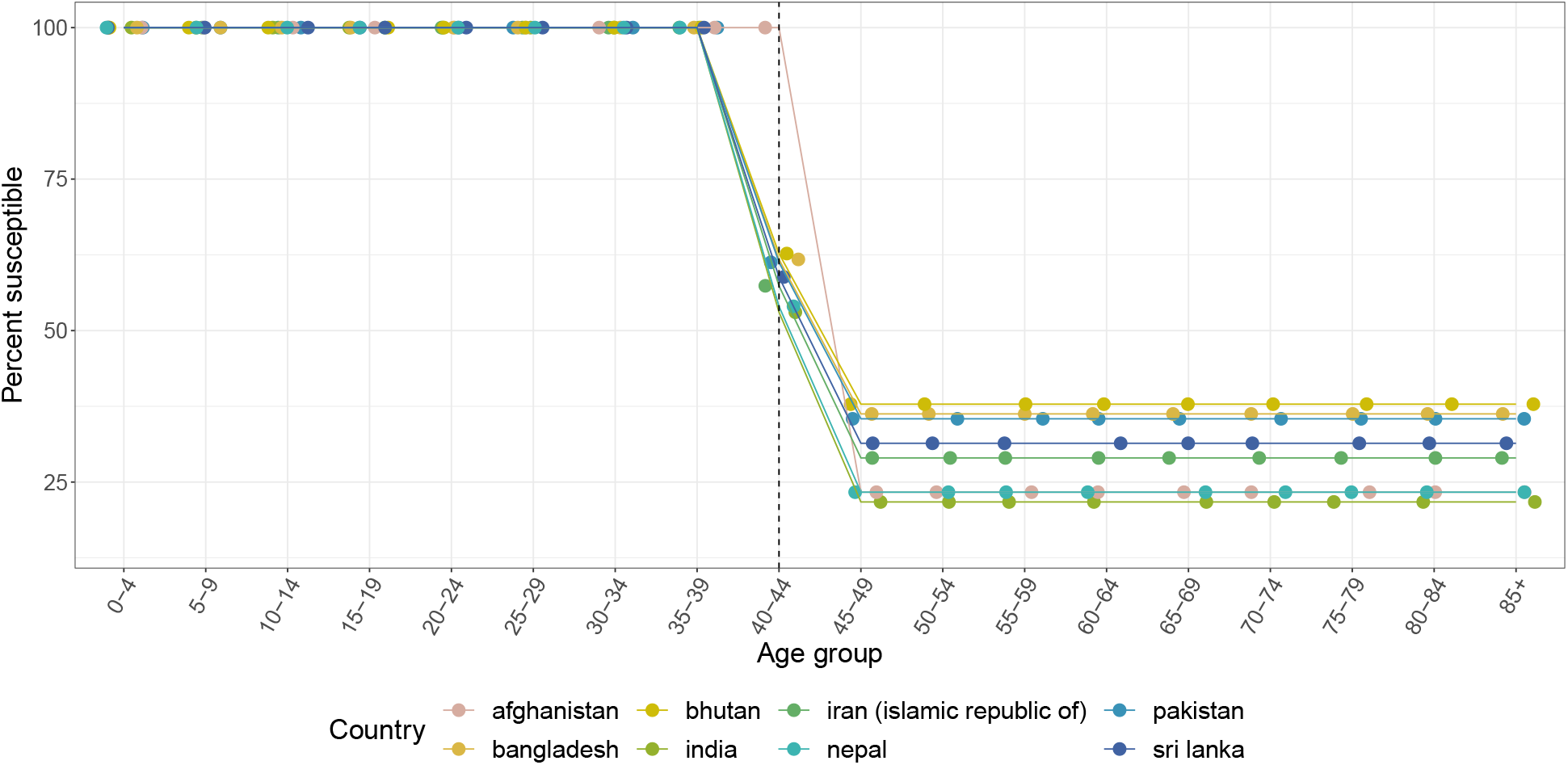
Country-specific monkeypox susceptibility profiles for countries in Central Asia. Susceptibility is calculated as 1 − (*ϵ* · `proportion vaccinated`) for each 5-year age group, where *ϵ* = 80 · 7% is the smallpox vaccine effectiveness against monkeypox. Dashed line indicates last age group in which some individuals were born before global smallpox eradication (1980). Points are jittered horizontally for visual aid.

**Figure S24:**
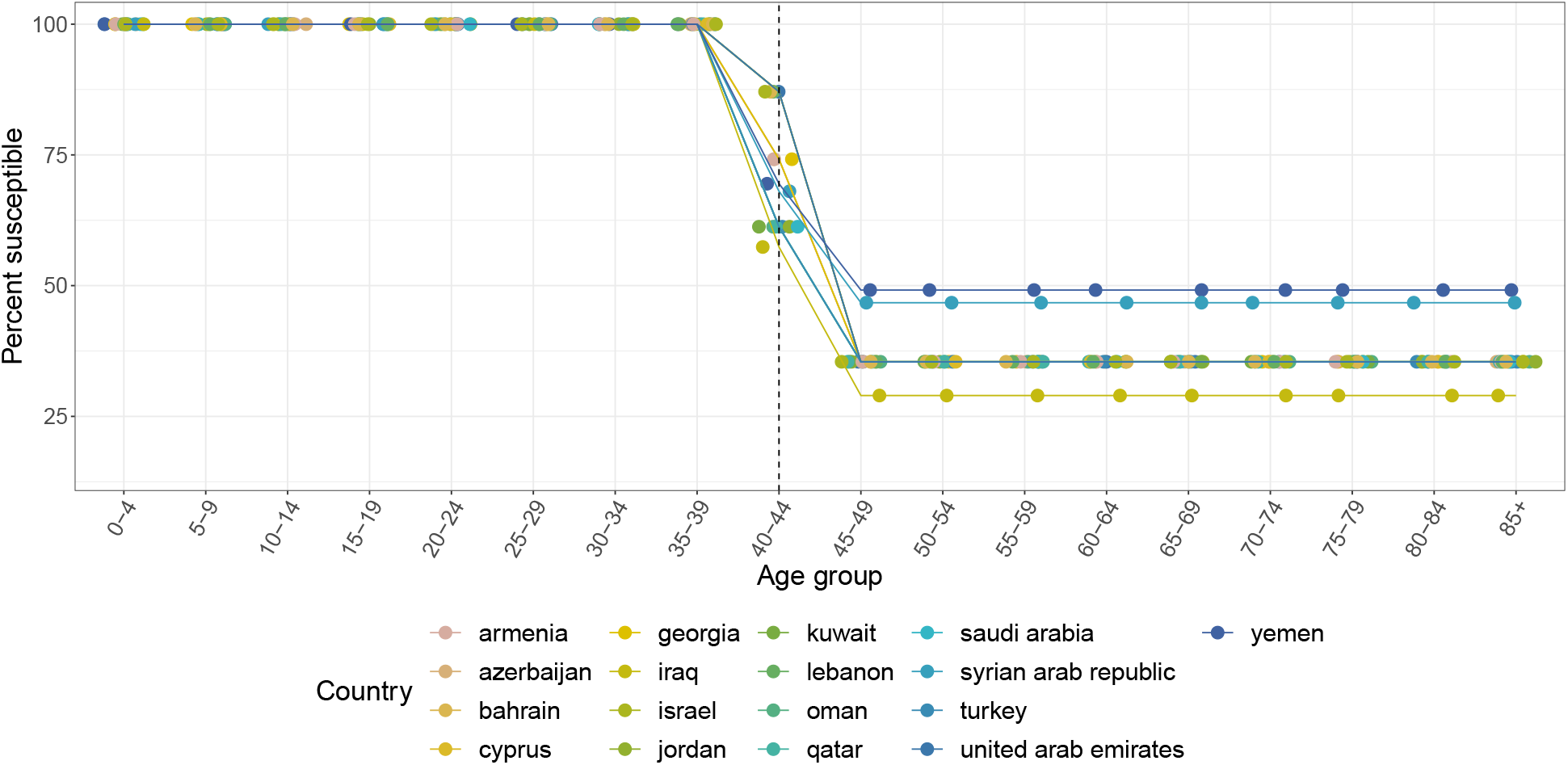
Country-specific monkeypox susceptibility profiles for countries in Western Asia. Susceptibility is calculated as 1 − (*ϵ* · `proportion vaccinated`) for each 5-year age group, where *ϵ* = 80 · 7% is the smallpox vaccine effectiveness against monkeypox. Dashed line indicates last age group in which some individuals were born before global smallpox eradication (1980). Points are jittered horizontally for visual aid.

**Figure S25:**
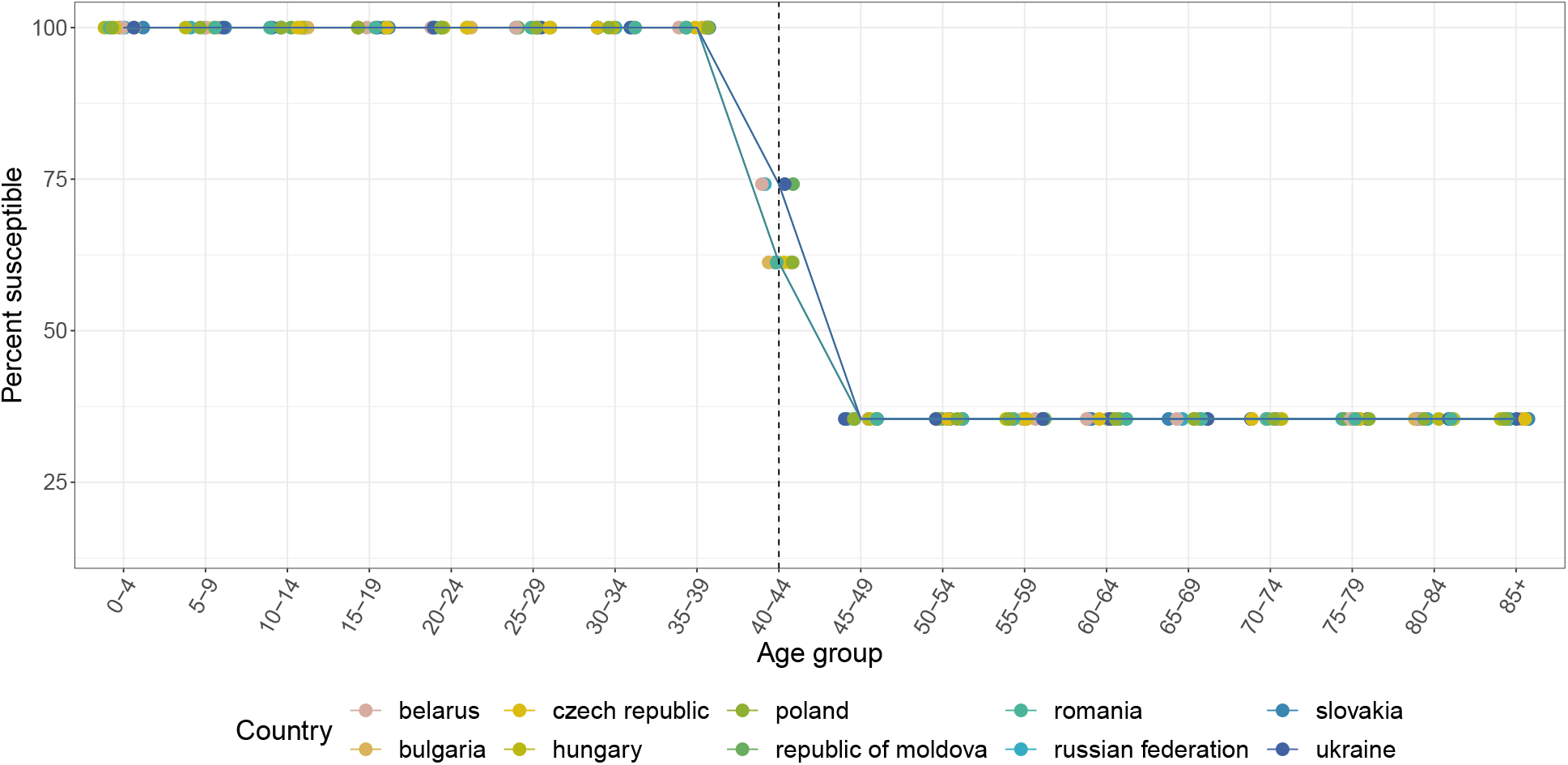
Country-specific monkeypox susceptibility profiles for countries in Eastern Europe. Susceptibility is calculated as 1 − (*ϵ* · `proportion vaccinated`) for each 5-year age group, where *ϵ* = 80 · 7% is the smallpox vaccine effectiveness against monkeypox. Dashed line indicates last age group in which some individuals were born before global smallpox eradication (1980). Points are jittered horizontally for visual aid.

**Figure S26:**
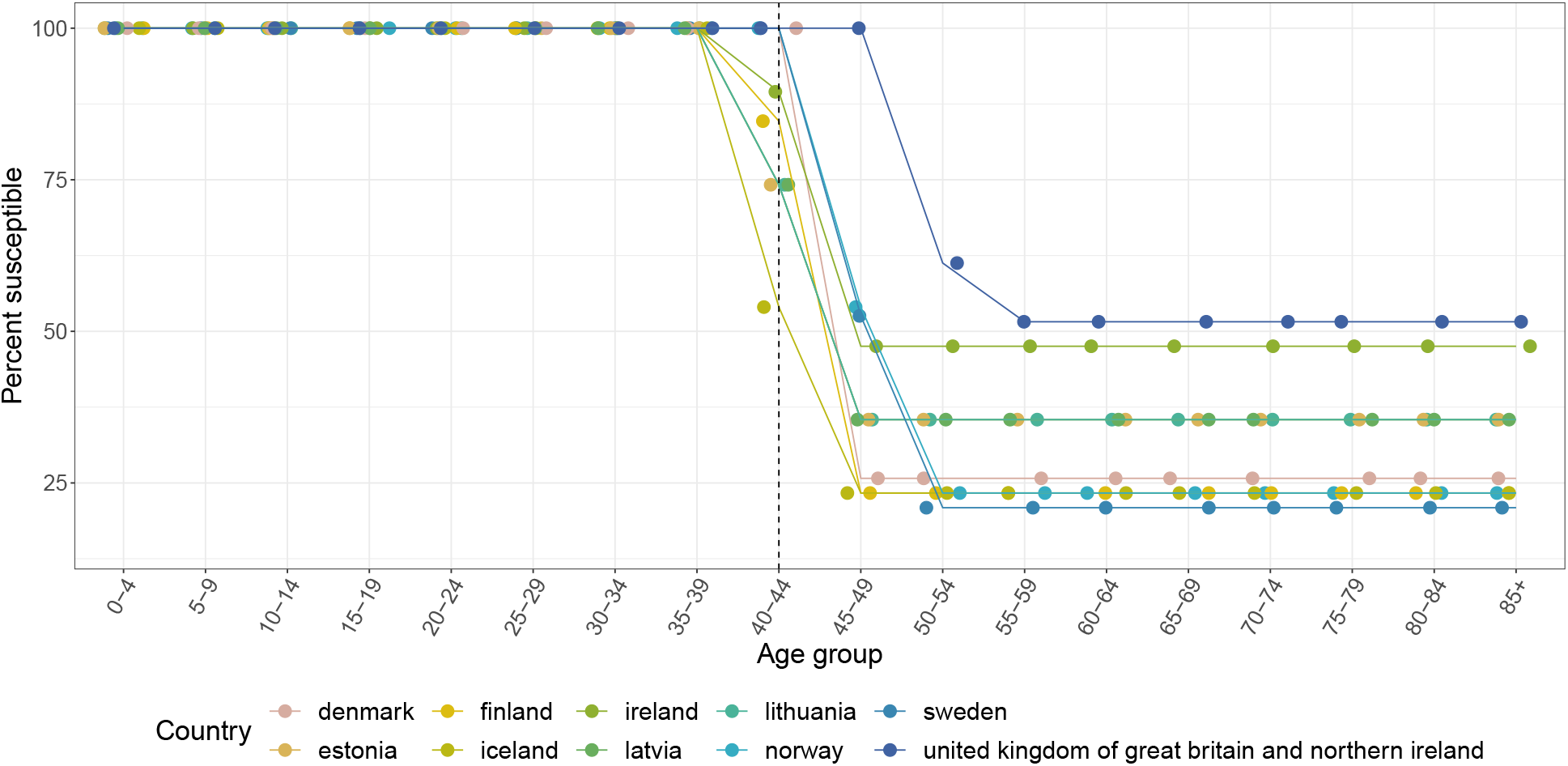
Country-specific monkeypox susceptibility profiles for countries in Northern Europe. Susceptibility is calculated as 1 − (*ϵ* · `proportion vaccinated`) for each 5-year age group, where *ϵ* = 80 · 7% is the smallpox vaccine effectiveness against monkeypox. Dashed line indicates last age group in which some individuals were born before global smallpox eradication (1980). Points are jittered horizontally

**Figure S27:**
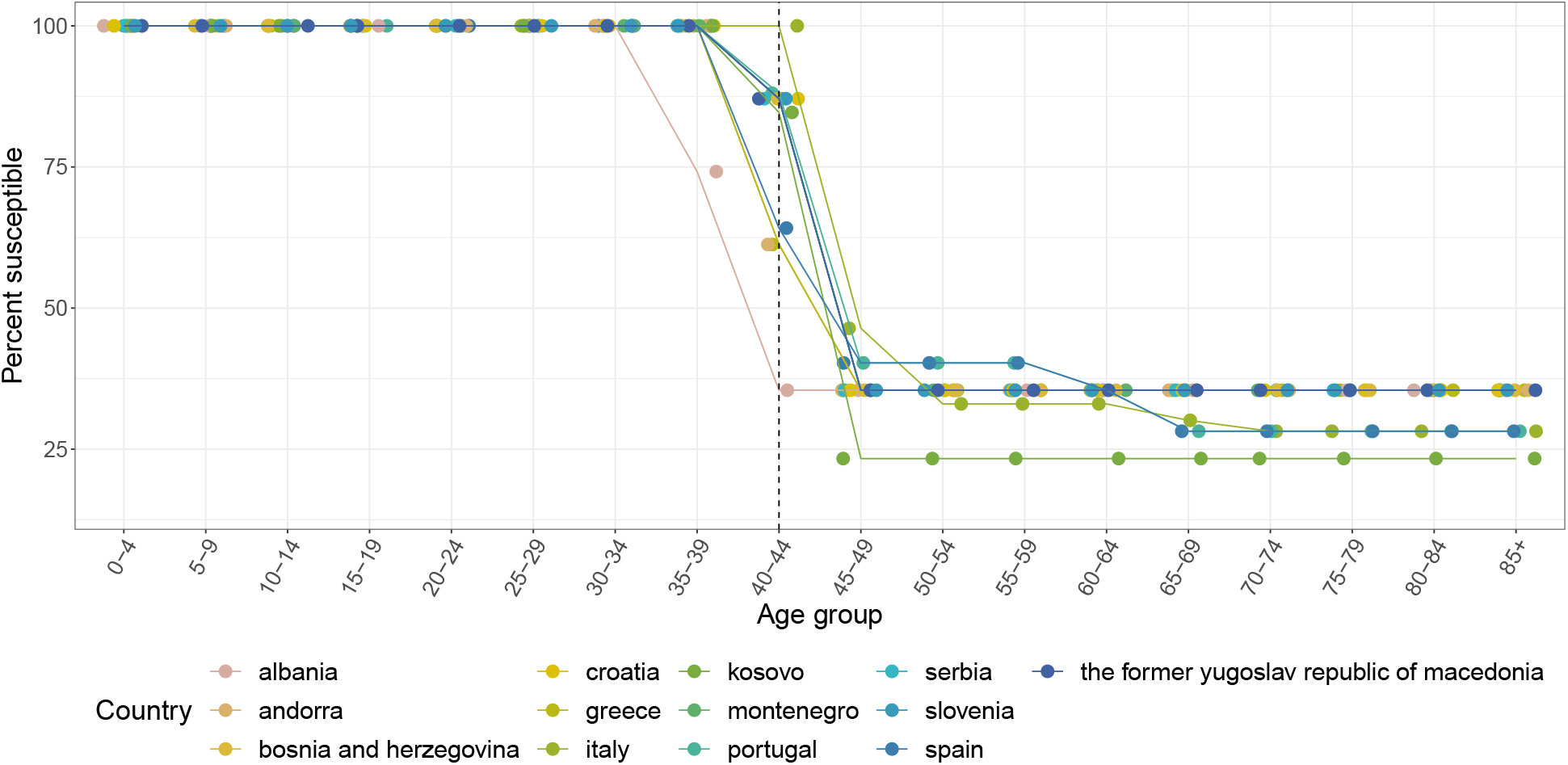
Country-specific monkeypox susceptibility profiles for countries in Southern Europe. Susceptibility is calculated as 1 − (*ϵ* · `proportion vaccinated`) for each 5-year age group, where *ϵ* = 80 · 7% is the smallpox vaccine effectiveness against monkeypox. Dashed line indicates last age group in which some individuals were born before global smallpox eradication (1980). Points are jittered horizontally for visual aid.

**Figure S28:**
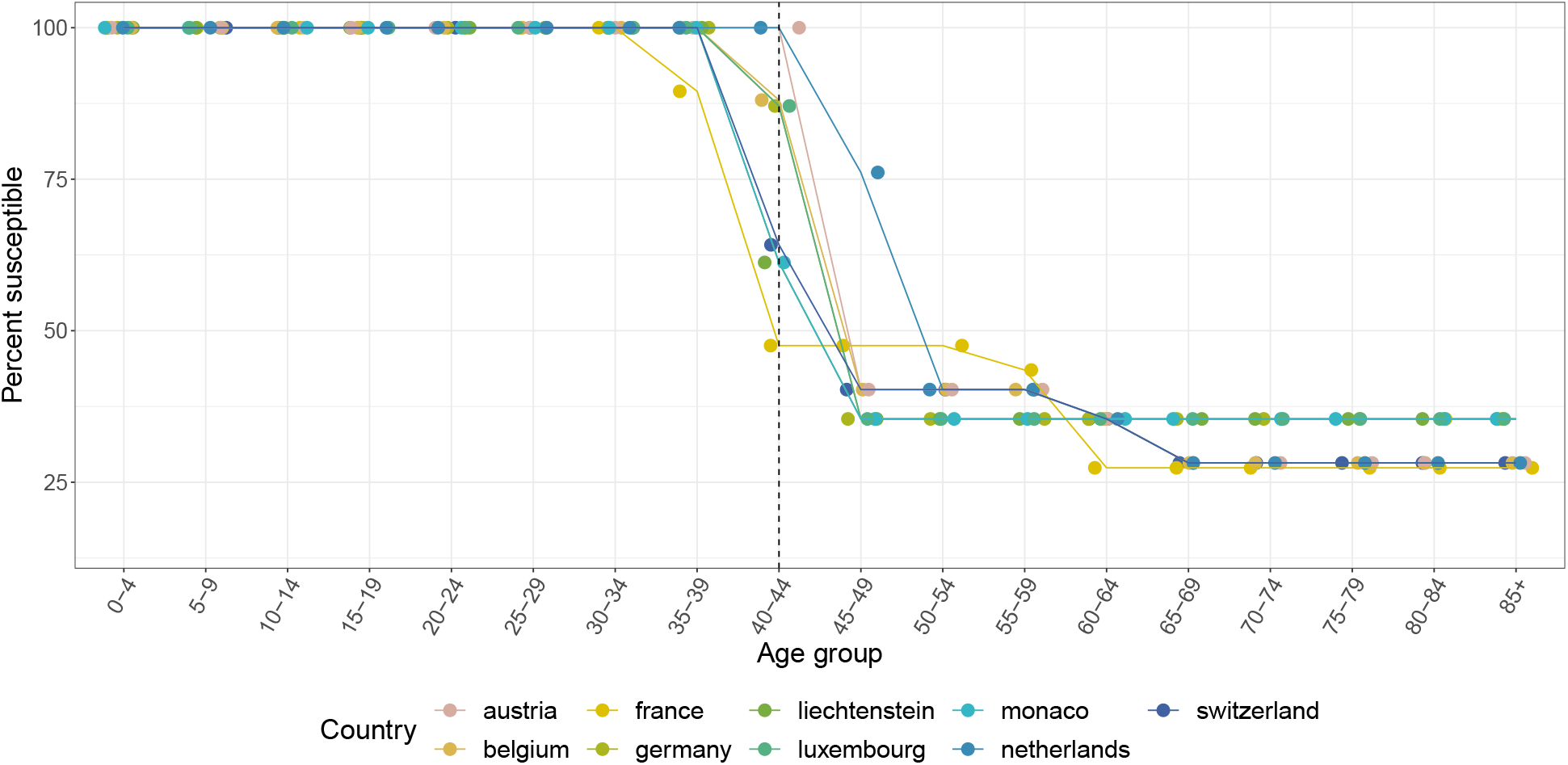
Country-specific monkeypox susceptibility profiles for countries in Western Europe. Susceptibility is calculated as 1 − (*ϵ* · `proportion vaccinated`) for each 5-year age group, where *ϵ* = 80 · 7% is the smallpox vaccine effectiveness against monkeypox. Dashed line indicates last age group in which some individuals were born before global smallpox eradication (1980). Points are jittered horizontally for visual aid.

**Figure S29:**
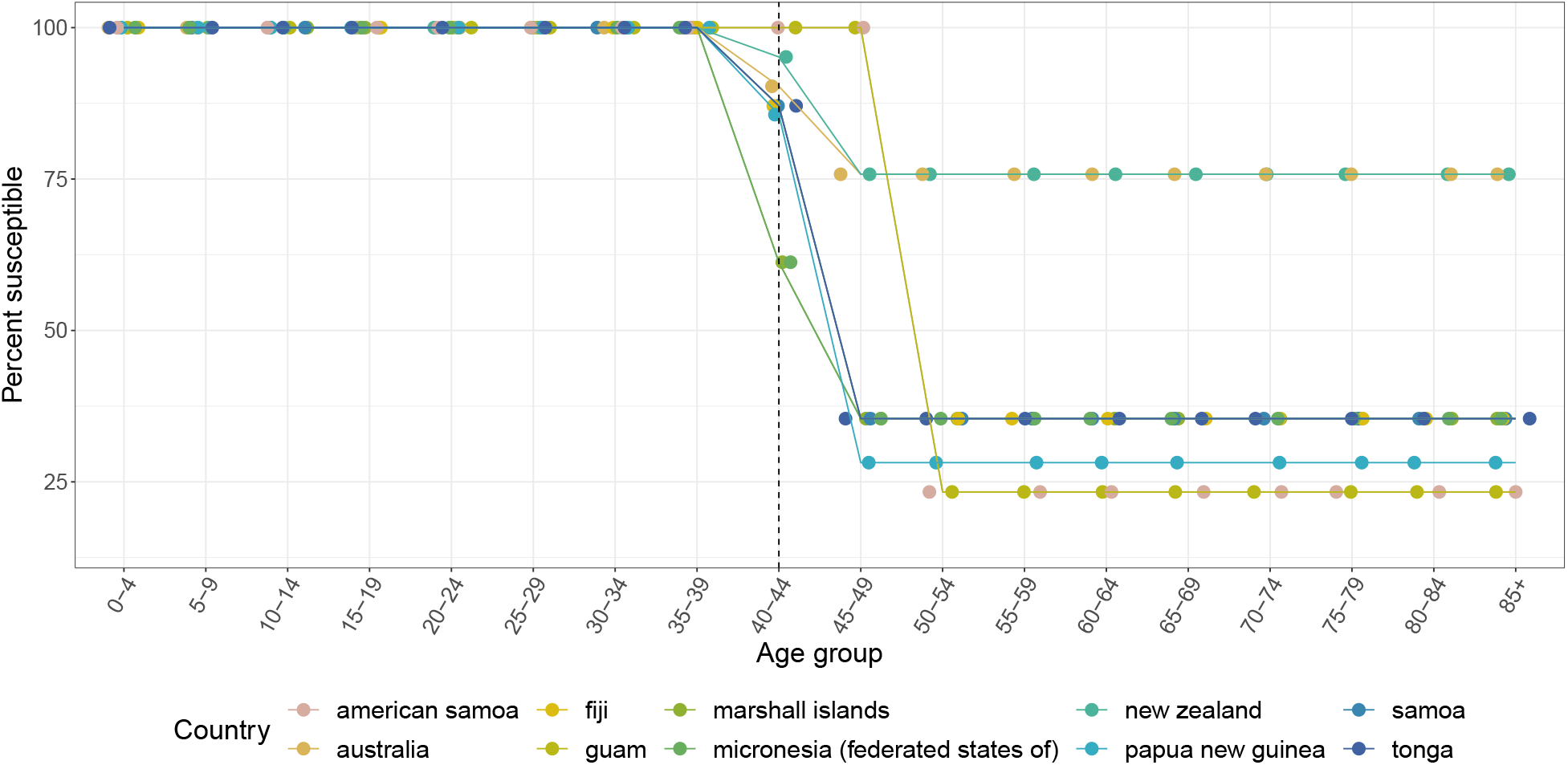
Country-specific monkeypox susceptibility profiles for countries in Oceania. Susceptibility is calculated as 1 − (*ϵ* · `proportion vaccinated`) for each 5-year age group, where *ϵ* = 80 · 7% is the smallpox vaccine effectiveness against monkeypox. Dashed line indicates last age group in which some individuals were born before global smallpox eradication (1980). Points are jittered horizontally for visual aid.

**Figure S30:**
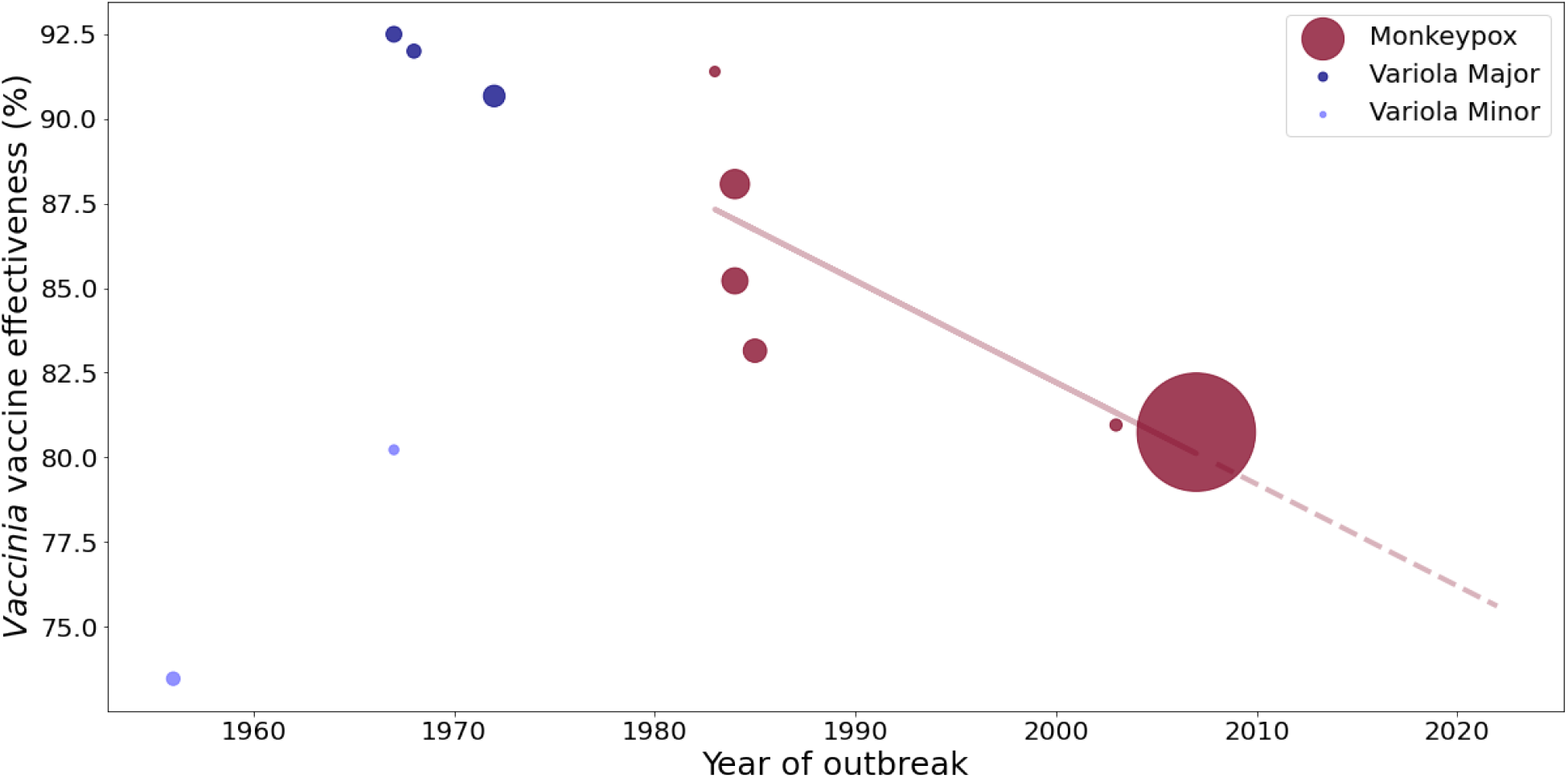
Smallpox (*Vaccinia*) vaccine effectiveness against orthopoxviruses. Data are from a literature search, and include studies that provided data on cases and non-cases by vaccination status. Additionally, studies after 1990 were only included if they differentiated smallpox-vaccine eligible (e.g., born before 1980) individuals. The marker size denotes study sample size. The line is based on a linear regression of all available monkeypox data, and the dashed portion is an extrapolation of this relationship to 2022.

